# Genome-wide association study meta-analysis identifies three novel loci for circulating anti-Müllerian hormone levels in women

**DOI:** 10.1101/2020.10.29.20221390

**Authors:** Renée MG Verdiesen, Yvonne T van der Schouw, Carla H van Gils, WM Monique Verschuren, Frank JM Broekmans, Maria C Borges, Ana LG Soares, Deborah A Lawlor, A Heather Eliassen, Peter Kraft, Dale P Sandler, Sioban D Harlow, Jennifer A Smith, Nanette Santoro, Minouk J Schoemaker, Anthony J Swerdlow, Anna Murray, Katherine S Ruth, N Charlotte Onland-Moret

## Abstract

Anti-Müllerian hormone (AMH) is expressed by antral stage ovarian follicles in women. Consequently, circulating AMH levels are detectable until menopause. Variation in age-specific AMH levels has been associated with breast cancer and polycystic ovary syndrome (PCOS), amongst other diseases. Identification of genetic variants underlying variation in AMH levels could provide clues about the physiological mechanisms that explain these AMH-disease associations. To date, only one variant in *MCM8* has been identified to be associated with circulating AMH levels in women. We aimed to identify additional variants for AMH through a GWAS meta-analysis including data from 7049 premenopausal women of European ancestry, which more than doubles the sample size of the largest previous GWAS. We identified four loci associated with AMH levels at p < 5×10^−8^: the previously reported *MCM8* locus and three novel signals in or near *AMH, TEX41*, and *CDCA7*. The strongest signal was a missense variant in the *AMH* gene (rs10417628). Most prioritized genes at the other three identified loci were involved in cell cycle regulation. Genetic correlation analyses indicated a strong positive correlation among SNPs for AMH levels and for age at menopause (r_g_= 0.82, FDR=0.003). Exploratory Mendelian randomization analyses did not support a causal effect of AMH on breast cancer or PCOS risk, but should be interpreted with caution as they may be underpowered and the validity of genetic instruments could not be extensively explored. In conclusion, we identified a variant in the *AMH* gene and three other loci that may affect circulating AMH levels in women.

## Introduction

Anti-Müllerian hormone (AMH) is generally known for its function in sexual differentiation, during which AMH signaling is essential for the regression of internal female reproductive organs in male embryos.^1^ In women, AMH is expressed by granulosa cells of primary ovarian follicles, and AMH expression continues until the antral stage.^2^ AMH becomes undetectable after menopause, when the ovarian reserve is depleted, and AMH can therefore be used as a marker for reproductive aging.^3^

Variation in age-specific circulating AMH levels has been associated with the occurrence of several non-communicable diseases, including breast cancer.^4^ In addition, it has been suggested that AMH may be involved in the pathogenesis of polycystic ovary syndrome (PCOS).^5^ Gaining more insight into genetic variation and biological mechanisms underlying inter-individual variation in AMH expression through genome-wide association studies (GWASs) could provide new clues regarding postnatal functions of AMH, and possibly, the etiologies of non-communicable diseases associated with AMH levels.

Previous GWASs on circulating AMH levels included either a mixture of male and female adolescents^6^, a very small study population^7^, or women of late reproductive age^8^ in whom AMH levels are generally very low. Of the previous GWASs, only the largest (n = 3344) identified a single genetic variant for AMH levels in premenopausal women, at chromosome 20 (rs16991615)^8^, which is also associated with natural age at menopause.^9; 10^ As sample sizes of previous GWASs were relatively small, a larger GWAS meta-analysis might lead to detection of more AMH variation loci. Moreover, as most variation in AMH levels in women is observed at ages 20 to 40 years^11^, including younger women will increase power to identify additional loci. Therefore, we aimed to identify additional genetic variants for AMH through a GWAS meta-analysis including 7049 premenopausal female participants. For that, we combined summary statistics from the AMH GWAS meta-analysis by Ruth et al.^8^ with GWAS data from 3705 additional women of early and middle reproductive age from 3 different cohorts.

## Subjects and Methods

### Study population

We included data from 7049 premenopausal female participants (median age ranged from 15.3 to 48 years across cohorts; Table 1) of European ancestry. In addition to the data from the AMH GWAS meta-analysis by Ruth et al.^8^ (n = 3344), we included data from the Doetinchem Cohort Study^12; 13^ (n = 2084), the Study of Women’s Health Across the Nation (SWAN)^14^ (n = 425), and data from adolescent daughters of the Avon Longitudinal Study of Parents and Children (ALSPAC)^15^ (n = 1196). The GWAS by Ruth et al. included data from the Generations Study^16^, Sister Study^17^, Nurses’ Health Study^18^, Nurses’ Health Study II^19^, and ALSPAC mothers^20^. For the current study, we requested summary statistics excluding data from ALSPAC mothers, as we wanted to analyze data from the ALSPAC mothers separately to investigate potential bias due to cryptic relatedness. More details about participating studies and the definitions used for the assessment of menopausal status are described in the Supplemental Methods and Supplemental Table 1. All studies received ethical approval from an institutional ethics committee.

**Table 1:** Distributions of AMH and age per participating study.

| **Study** | **N** | **AMH, pmol/L**<br>**(median (IQR))** | **Age at blood collection, years**<br>**(median (IQR))** |
| --- | --- | --- | --- |
| Studies contributing to summary statistics GWAS Ruth et al.<sup>8*</sup> |  |  |  |
| Generations Study | 379 | 3.9 (0.8, 11.7) | 44 (40, 48) |
| Sister Study | 438 | 1.2 (0.1, 6.0) | 48 (45, 51) |
| Nurses' Health Studies | 642 | 6.1 (2.0, 13.9) | 44 (41, 47) |
| Additional studies |  |  |  |
| Doetinchem Cohort Study | 2,084 | 10.9 (2.9, 25.6) | 37.2 (31.2, 42.9) |
| ALSPAC mothers | 1,885 | 2.0 (0.4, 5.2) | 46 (44, 49) |
| ALSPAC daughters | 1,196 | 26.1 (18.2, 39.8) | 15.3 (15.3, 15.5) |
| SWAN | 425 | 1.1 (0.2, 3.3) | 47.3 (45.3, 49.3) |
| **Total** | **7,049** |  |  |

\* In the original study ALSPAC mothers were included as well, but in the current analyses summary statistics from ALSPAC were included separately to assess potential genomic inflation due to inclusion of both ALSPAC mothers and daughters. Therefore, we treated the ALSPAC mothers as individual study.

### AMH measurements

Included studies measured AMH in either serum or plasma using different AMH ELISA assays. Also, the methodology for handling AMH measurements below the assay limit of detection (LOD) differed across studies. A detailed overview of these study-specific details has been included in Supplemental Table 1. Across studies, the percentage of measurements under the assay-specific LODs ranged from 0% to 24.2%.

### Genotyping and imputation

Extensive details on genotyping and imputation procedures for each participating study are presented in Supplemental Table 2. Briefly, samples of the Generations Study, Sister Study, and most samples of the Nurses’ Health Studies, were genotyped using the OncoArray array.^8^ The remaining 225 samples of the Nurses’ Health Studies were genotyped using Illumina HumanHap550 and HumanHap610 arrays.^8^ Samples of the Doetinchem Cohort Study were genotyped using the Illumina Infinium Global Screening Array-24 Kit (Illumina Inc., San Diego, California, United States of America). For genotyping of samples from ALSPAC mothers and daughters, the Illumina Human660W-Quad array and Illumina HumanHap550 quad genome-wide SNP genotyping platform were used, respectively. SWAN participants were genotyped using the Illumina Multi-Ethnic Global Array (MEGA A1). All participating studies performed sample and SNP QC prior to imputation, which was done using the Haplotype Reference Consortium (HRC) panel version r1.1 2016 (Supplemental Table 2).

### Association analyses

All studies converted AMH concentrations to pmol/L using 1 pg/mL = 0.00714 pmol/L. As AMH levels are not normally distributed, AMH measurements were transformed using rank-based inverse normal transformation in all studies, as previously described.^8^

In all studies linear models were fitted, assuming additive SNP effects, adjusted for age at blood collection (years) (Supplemental Methods). Analyses were further adjusted for population stratification by inclusion of either 10 principal components (ALSPAC, SWAN) or a kinship matrix (Generations Study, Sister Study, Nurses’ Health Studies, Doetinchem Cohort Study). In addition, we included summary statistics of the meta-analysis of the Generations Study, Sister Study, and Nurses’ Health Studies, which was performed using METAL, as described elsewhere.^8^ Separate association analyses were conducted for the ALSPAC mothers and daughters, because of the large differences in both age and AMH distributions between these groups (Supplemental Methods).

Prior to meta-analysis, we performed file-level and meta-level QC on all summary statistics files to clean and check the data, and to identify potential study-specific problems. File-level and meta-level QC were conducted using the R package EasyQC (v9.2), following a previously published protocol^21^ (Supplemental Methods). No study-specific issues were identified through these QC procedures (Figure S1-S5). In addition, we sought to confirm that inclusion of ALSPAC mothers and daughters as separate cohorts would not result in inflation of effect estimates due to cryptic relatedness (411 mother-daughter pairs were present in the ALSPAC data). We checked this through meta-analyzing only data of the two ALSPAC cohorts and checking both the corresponding QQ plot and calculating λ. Given the absence of genomic inflation (λ = 1.01, QQplot in Figure S6) we included summary statistics of both ALSPAC mothers and daughters in the meta-analysis.

We performed an inverse variance weighted meta-analysis using METAL (version 2011-03-25). Genomic control was applied for all included studies. SNPs with a minor allele frequency (MAF) < 1% and/or poor imputation quality (info score < 0.4 or r^2^ < 0.3, depending on which metric was provided) were excluded. As a result, 8,298,138 autosomal SNPs were included in this AMH GWAS meta-analysis. To assess if observed effect estimates were homogeneous across studies, we additionally performed a heterogeneity analysis in METAL.

To identify lead and secondary SNPs within genome-wide significant associated loci, we performed an approximate conditional and joint association analysis.^22^ We used Genome-wide Complex Trait Analysis (GCTA)^23^ (version 1.93.1f beta) to run a stepwise model selection procedure to select independently associated SNPs (cojo-slct) using the summary-level data. We estimated LD between SNPs using data of 4059 unrelated participants from the EPIC-NL cohort^24^ as LD reference panel.

Because of the strong correlation between AMH and age, and the difference in both the AMH and age distributions in the ALSPAC daughters compared to the other included cohorts, we performed a sensitivity analysis in which we excluded the ALSPAC daughters. Furthermore, this sensitivity analysis served as an additional check that inclusion of both ALSPAC mothers and daughters did not cause identification of false-positive hits.

### Gene-based genome-wide association analysis

We performed a gene-based genome-wide association analysis using the MAGMA^25^ implementation (v1.07) in the online Functional Mapping and Annotation of Genome-wide Association Studies (FUMA) platform (FUMA)^26^ (parameter settings are listed in Table S3). For this analysis, SNPs located in gene bodies were aggregated to 18,896 protein coding genes (Ensembl build 92). MAGMA tests the joint association of all SNPs in each gene with inverse normal transformed AMH levels using a multiple linear regression approach, which takes LD between SNPs into account.^25^ FUMA considered genes to be significantly associated with circulating AMH levels if p < 2.65 × 10^−6^ (Bonferroni corrected p-value; 0.05/ 18,896).

### Functional annotation using FUMA

FUMA is an integrative web-based platform that uses 18 biological resources and can be used to functionally annotate lead variants from GWAS, and to prioritize the most likely causal SNPs and genes^26^. We used the SNP2GENE process integrated into FUMA (v1.3.6)^26^ for the characterization of genomic loci and functional gene mapping (parameter settings are listed in Table S3). We included SNPs identified in our approximate conditional and joint analysis as predefined lead SNPs for the characterization of genomic risk loci. SNPs that were in LD with these lead SNPs (r^2^ > 0.6) within a 500kb window based on the 1000G Phase 3 European reference panel population in FUMA, and a GWAS meta-analysis p-value < 0.05 were selected as candidate SNPs. Non-GWAS-tagged SNPs from the 1000G Phase 3 European reference that met these LD and distance criteria were also selected as candidate SNPs. Candidate SNPs were annotated based on Combined Annotation Dependent Depletion (CADD) scores^27^, Regulome DB scores^28^, and chromatin states^29^ (Table S3). Positional mapping, eQTL mapping and chromatin interaction mapping were used to map SNPs to genes (Table S3). For chromatin states, eQTL mapping and chromatin interaction mapping we only selected tissues and cell types that are most likely to be involved in AMH expression and signaling (Table S3).

### Pathway analysis using DEPICT

We used the hypothesis-free pathway analysis tool DEPICT (v1)^30^ to prioritize the most likely causal genes at associated loci, to highlight gene sets enriched in genes within associated loci, and to identify tissues/cell types that are implicated by the associated loci. For these analyses, we included all suggestive significant SNPs (p < 5 × 10^−6^), which were clumped at LD r^2^ < 0.1 and a physical distance of 500kb using PLINK v.1.9 as part of the DEPICT pipeline.

### LD Score Regression

We estimated SNP heritability using the LD Hub web interface (v1.9.3)^31^ for LD score regression^32^. In addition, we used LD Hub to estimate SNP-based genetic correlations between AMH and phenotypes that have been associated with AMH in observational studies. These genetic correlation analyses make use of GWAS summary statistics for all SNPs to estimate genetic covariance among SNPs for two traits.^33^ Included phenotypes comprised reproductive traits, hormones (leptin), anthropometric traits, blood lipids, glycemic traits, metabolites, cardiometabolic traits, cancer, autoimmune diseases, bone mineral density, aging and smoking behavior. Of the 597 UK Biobank traits in the LDHub database, we only included traits that corresponded to these phenotype categories, resulting in 345 comparisons. To correct for multiple testing, we calculated false discovery rates (FDR), using the p.adjust function in R (R package “stats”).^34^ FDR adjusted p-values < 0.05 were considered to be significant.

### Mendelian randomization

In observational studies, AMH has been associated with breast cancer^4^, and PCOS^35^, amongst other diseases. As the exact function of AMH in the etiology of these diseases is unclear, and actual AMH levels are associated with predicted future age at menopause and current menopausal status, causality of these AMH-disease associations remains to be determined. Mendelian randomization (MR) is a method that may provide evidence for causality of observational associations.^36^ Because our AMH GWAS meta-analysis only included women, and previous research suggests that genetic variants for inter-individual differences in AMH levels differ between males and females^6^, we performed MR analyses for the female-specific outcomes breast cancer and PCOS only. We performed two-sample MR analyses^37^ using the R package TwoSampleMR (version 0.5.1)^38^. We included identified lead SNPs as genetic instruments for AMH. For the outcomes, we included summary statistics from the most recent largest GWASs for breast cancer (n = 228,951; 122,977 cases)^39^ and PCOS (n = 113,238; 10,074 cases)^40^. Wald ratio estimates were calculated for individual SNPs and a random effects inverse variance weighted (IVW) meta-analysis approach was used to combine these estimates. To assess the strength of included genetic variants for AMH we calculated F-statistics corresponding to the IVW analyses, using the proportion of variance in AMH explained by the genetic variants, the sample size of the outcome GWASs, and the number of variants included^41^. We compared the overall MR estimate (i.e. IVW) to SNP-specific MR estimates (i.e. Wald ratio) since inconsistent estimates are indicative of horizontal pleiotropy, which is a violation of the MR assumptions.^38^ In addition, we tested for heterogeneity in causal effects amongst the genetic instruments using Cochrane’s Q statistics and performed leave-one-out sensitivity analyses to assess the potential effect of outlying variants.

## Results

Descriptive statistics on age and AMH levels of the study participants included in this GWAS meta-analysis are presented per study in Table 1. Median AMH ranged from 1.1 pmol/L in SWAN to 26.1 pmol/L in ALSPAC daughters. Median age ranged from 15.3 years in ALSPAC daughters to 48 years in the Sister Study.

### Genome-wide association analysis

We identified four genome-wide significant lead SNPs (p <5 × 10^−8^) for inverse normally transformed AMH, in four loci (Table 2, Figure 1, Figure S7-8). Approximate conditional and joint analysis did not reveal secondary signals. In addition to the previously reported locus on chromosome 20 (rs16991615, nearest gene: *MCM8*), we identified 1 locus on chromosome 19 (nearest gene: *AMH*) and 2 loci on chromosome 2 (nearest genes: *TEX41* and *CDCA7*). The strongest signal was rs10417628 on chromosome 19, which is physically located in the *AMH* gene (β = −0.34, se = 0.05, p = 1.2 × 10^−11^) (Figure S8A). Combined the four lead SNPs explained 1.47% of the variance in AMH levels. In the sensitivity analysis excluding ALSPAC daughters, all four loci from the main analysis remained genome-wide significant, and an additional locus at chromosome 5 (rs116090962, nearest gene: *CTB-99A3*.*1*) was identified (β = 0.38, se = 0.07, p = 6.0 × 10^−9^) (Table S4).

**Table 2:**
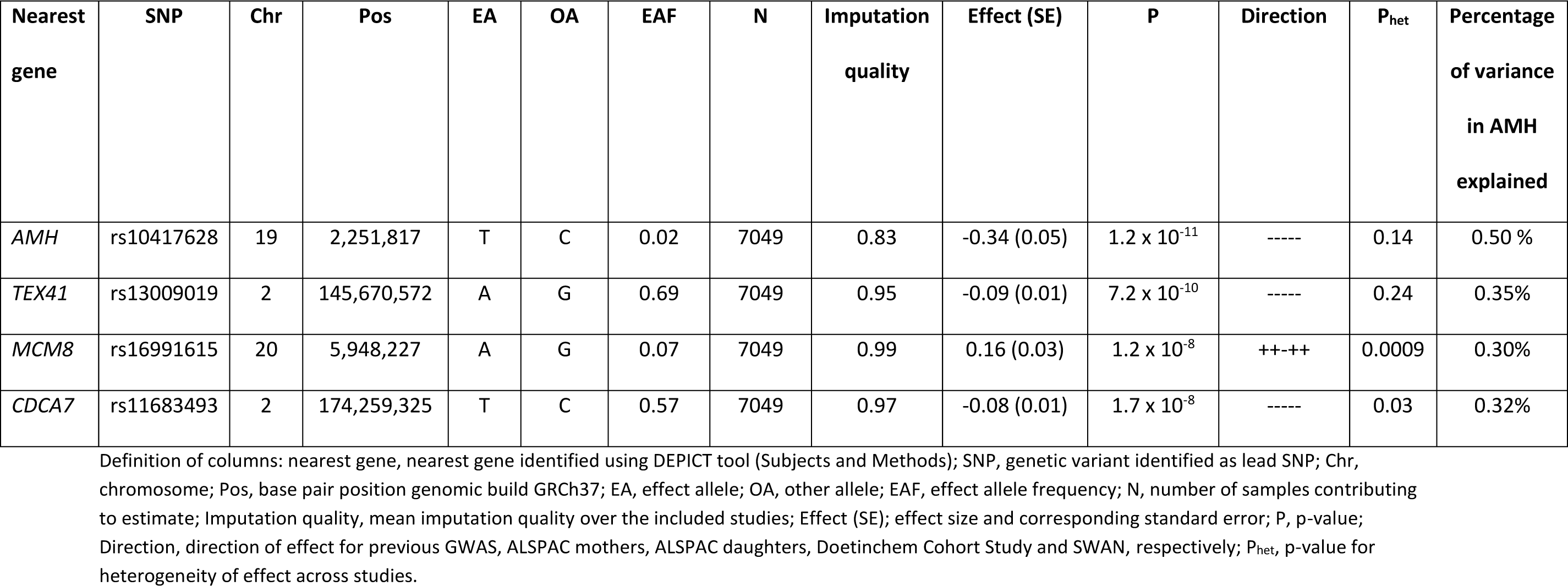
Loci significantly associated (p < 5 × 10^−8^) with inverse normally transformed AMH in women.

**Figure 1:**
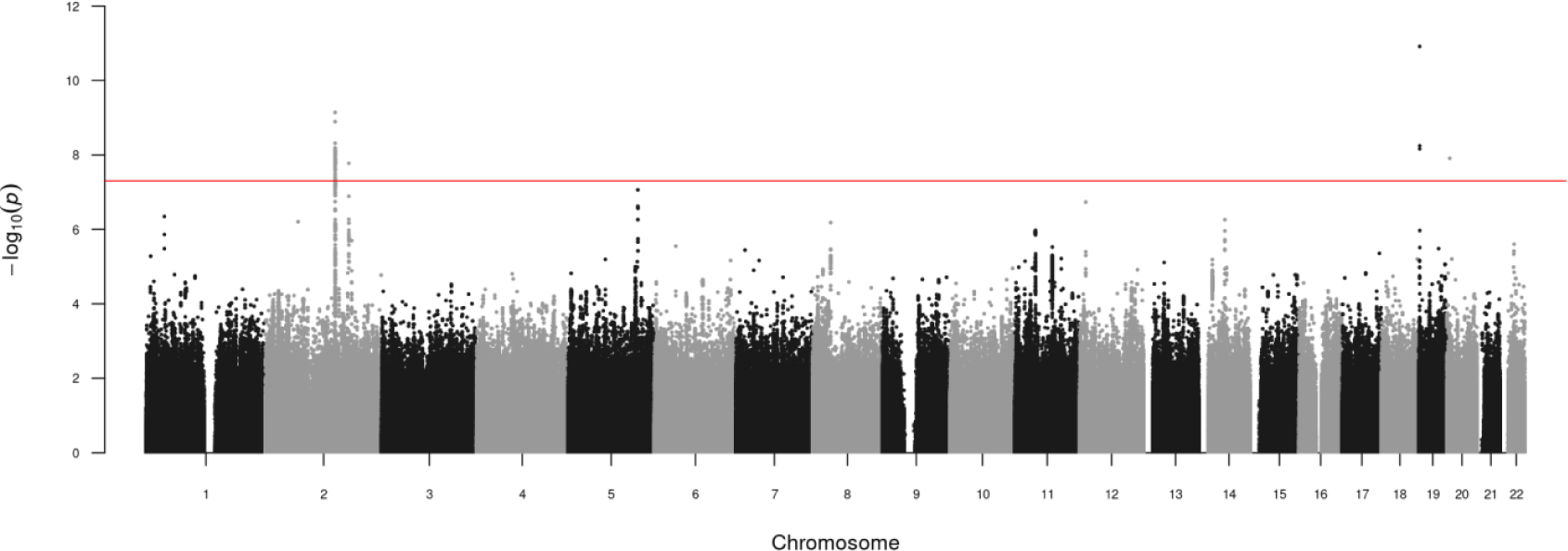
Manhattan plot of genome-wide association results for inverse normally transformed AMH in women. Results are association results from meta-analysis of inverse normally transformed AMH in 7,049 women of European ancestry. Individual studies adjusted analyses for age at AMH measurement and population stratification (through kinship matrix or 10 principal components).

### Gene-based genome-wide association analysis

Gene-based genome-wide association analysis, which tested associations between 18,896 protein coding genes and inverse normal transformed AMH, highlighted the following four significant genes: *AMH, BMP4, EIF4EBP1, and GAB2* (Supplemental Figure 9).

### Functional Annotation using FUMA

Through the SNP2GENE process, FUMA identified 82 candidate SNPs that were in LD with the four identified lead SNPs (Supplemental Table 5). These candidate SNPs were used for the prioritization of genes.

In total, 12 genes were mapped to the locus of the previously identified SNP on chromosome 20 (rs16991615) (Supplemental Table 6), of which *MCM8* and *CRLS1* were prioritized based on eQTL mapping (Figure 2A). *CRLS1* was the only gene prioritized based on both eQTL mapping and chromatin interactions. However, as rs16991615 is a missense variant located in exon 9 of the *MCM8* gene, and this was the only SNP identified for this locus, *MCM8* is the most likely gene causing this signal.

**Figure 2:**
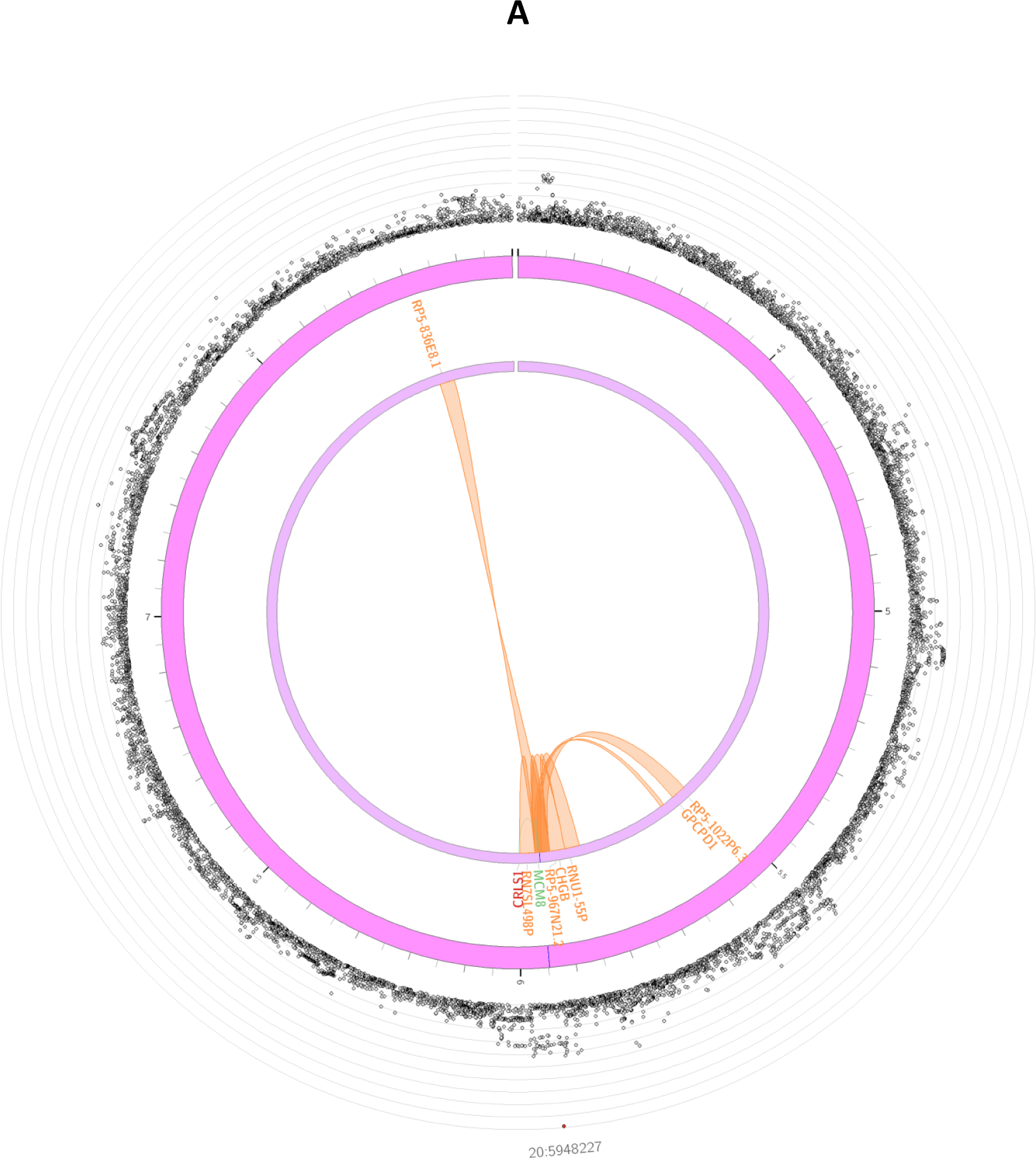
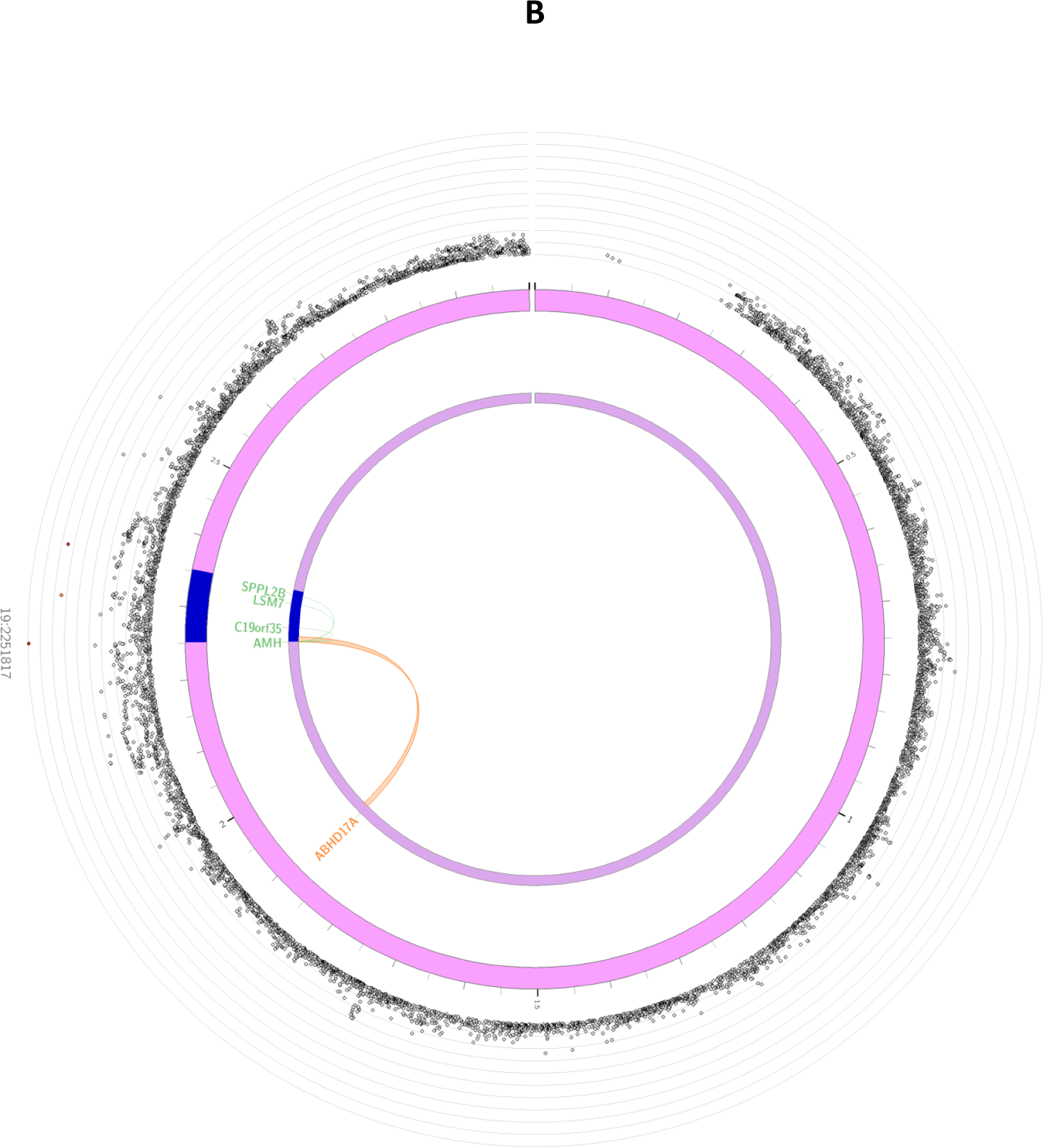
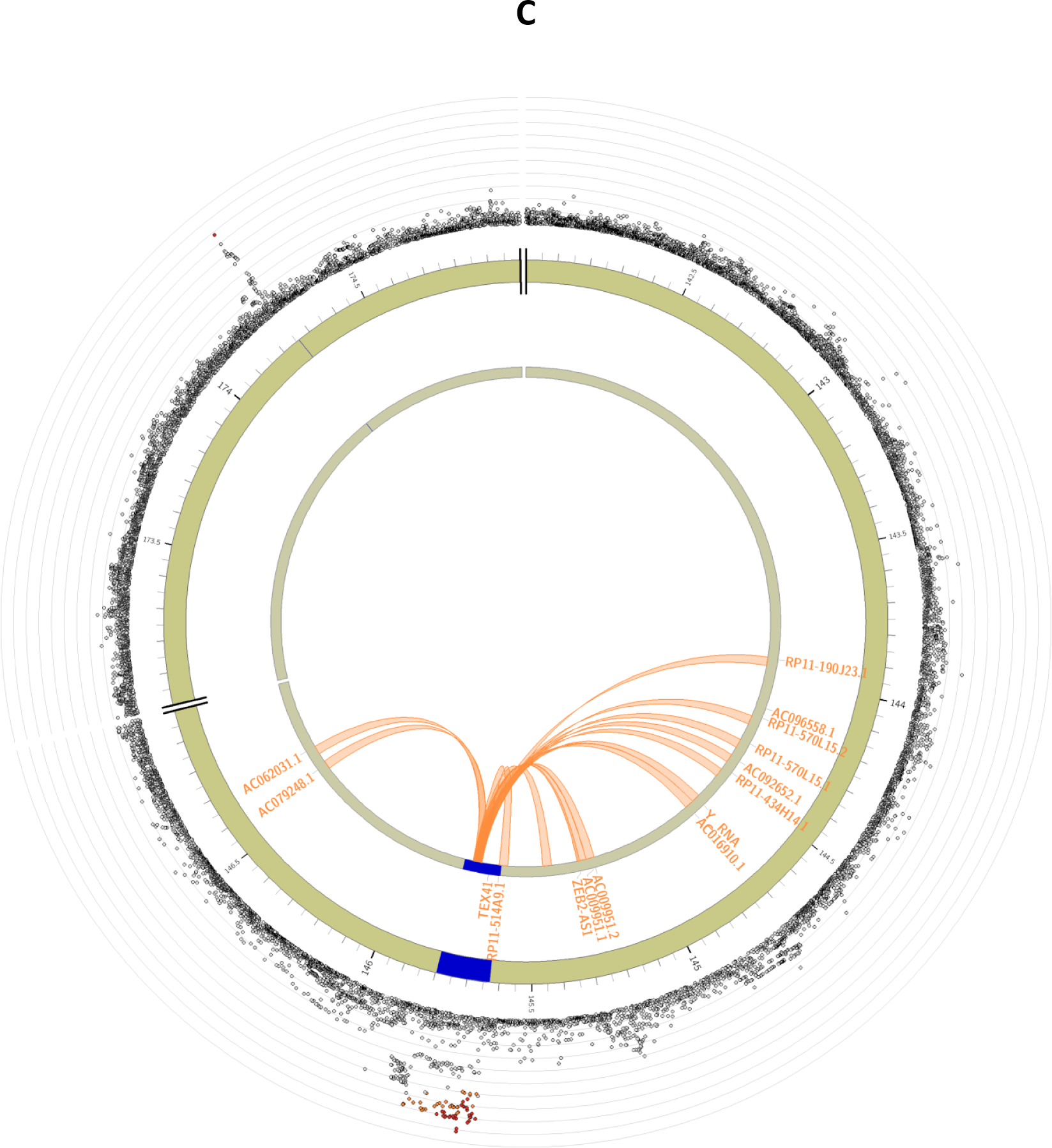
Circos plots for the genome-wide significant loci for inverse normally transformed AMH in women. Circos plots are presented for each of the identified loci: *MCM8* (panel A), *AMH* (panel B), *TEX41* and *CDCA7* (panel C). The outer layer represents the Manhattan plot. The second (including genomic positions) and third layers represents the chromosome ring, genomic risk loci are depicted in blue. Only genes mapped by either eQTLs or chromatin mapping are plotted. Genes only mapped by eQTLs are green, genes only mapped by chromatin interactions are orange, and genes mapped by both have a red colour. Orange coloured lines represent chromatin interactions, green coloured lines are eQTL links. Plots were created using the FUMA platform^26^.

For the locus on chromosome 19, 3 candidate SNPs (rs10417628, rs12462821, rs7247495) were identified. The lead SNP in this locus (rs10417628) is also a missense variant, located in exon 5 of the *AMH* gene, making this the most likely causal gene at this locus. The other 2 variants were located in intronic and intronic non-coding RNA regions. Based on the used parameter settings, FUMA mapped 8 genes to the *AMH* locus (Supplemental Table 6), of which 4 were highlighted by eQTL mapping (*AMH, C19orf35, SPPL2B and LSM7*) and 1 through chromatin interactions (*ABHD17A*) (Figure 2B).

Most of the candidate SNPs were identified for the locus on chromosome 2 near *TEX41*. All 77 variants were located in either intronic or exonic long noncoding RNA regions. Of the 15 genes mapped to this locus (Supplemental Table 6), no genes were prioritized based on eQTL mapping, but several genes were prioritized based on chromosome interactions, including *ZEB2-AS1* (Figure 2C). In the other locus on chromosome 2, for which *CDCA7* is the nearest gene, no additional candidate SNPs were identified and no genes were mapped to this locus.

### Pathway analysis using DEPICT

Using the DEPICT tool, 188 suggestive associated SNPs (p < 5 × 10^−6^) were clumped at LD r^2^ < 0.1 and a physical distance of 500 kb, resulting in 24 clumps as input for the enrichment analyses (Table S7). The top three prioritized gene sets were “URI1 PPI subnetwork”, “NFYB PPI subnetwork” and “nuclear inner membrane” (Table S8). “Induced Pluripotent Stem Cells” was identified as the highest prioritized cell type (Table S9). However, none of these enrichments were statistically significant (FDR > 0.05).

DEPICT prioritized nine genes at FDR < 0.05 as most likely causal genes (Table S10). Of the genome-wide associated loci, only *MCM8* (rs16991615), and *CDCA7* (rs11683493) were prioritized at this FDR threshold. *AMH, BMP4, and GAB2* were also prioritized by DEPICT, but FDR values were > 0.20. Of the genes significant in the gene-based MAGMA analysis, *EIF4EBP1* was prioritized at FDR < 0.05.

### LD Score Regression

We used LD score regression implemented in LD Hub to calculate SNP heritability for AMH based on the meta-analysis summary statistics. Total SNP heritability (h_g_^2^) on the observed scale was estimated to be 15% (se = 7%). We additionally performed genetic correlations analyses between AMH and 345 traits on LD Hub. After correction for multiple testing, AMH was only significantly correlated with age at menopause (r_g_ = 0.82, se = 0.19, FDR = 0.003) (Table S11).

### MR analyses

IVW MR estimates did not indicate a causal effect of circulating AMH on breast cancer risk (OR_IVW_ = 1.00, 95%CI: 0.74 – 1.36; Table 3). Results from the single SNP analysis including the variant in the *AMH* locus also did not support a causal association with breast cancer (OR_IVW_ = 0.99, 95%CI = 0.87 – 1.12), whereas analyses for the remaining variants suggested a risk decreasing effect of the SNPs in the *TEX41* and *CDCA7* loci and a risk increasing effect of the variant in the *MCM8* locus (Table 3). In agreement with these findings, a formal heterogeneity test for the IVW estimate indicated heterogeneity in causal effects amongst the four genetic variants (2.13 × 10^−11^), although the interpretation of this heterogeneity p-value is limited due to the small number of included SNPs. Leave-one-out sensitivity analyses supported the outlying effect of rs16991615 (*MCM8* locus) (Figure S11).

**Table 3:** Mendelian randomization estimates for causal effects of circulating AMH on breast cancer and PCOS risk.

| Outcome | Method | Odds Ratio | 95% CI | p |
| --- | --- | --- | --- | --- |
| Breast Cancer | IVW | 1.00 | 0.74 - 1.36 | 0.98 |
|  | Wald ratio estimate for rs10417628 (AMH) | 0.99 | 0.87 - 1.12 | 0.85 |
|  | Wald ratio estimate for rs13009019 (TEX41) | 0.84 | 0.72 - 0.97 | 0.02 |
|  | Wald ratio estimate for rs16991615 (MCM8) | 1.60 | 1.37 - 1.87 | 1.79 x 10<sup>-9</sup> |
|  | Wald ratio estimate for rs11683493 (CDCA7) | 0.76 | 0.65 - 0.89 | 9.41 x 10<sup>-4</sup> |
| PCOS | IVW | 1.29 | 0.85 - 1.95 | 0.23 |
|  | Wald ratio estimate for rs10417628 (AMH) | 1.27 | 0.64 - 2.56 | 0.49 |
|  | Wald ratio estimate for rs13009019 (TEX41) | 1.66 | 0.80 - 3.45 | 0.18 |
|  | Wald ratio estimate for rs16991615 (MCM8) | 1.75 | 0.83 - 3.69 | 0.14 |
|  | Wald ratio estimate for rs11683493 (CDCA7) | 0.66 | 0.29 - 1.50 | 0.32 |

Odds ratio and 95%CI are per 1 unit increase in inverse normally transformed AMH.
AMH, anti-Müllerian hormone; PCOS, polycystic ovary syndrome; IVW, inverse variance weighted; MR, Mendelian randomization.

For PCOS, the IVW MR estimate suggested that higher genetically predicted AMH levels are potentially associated with an increased risk of PCOS, but confidence intervals were wide and included the null (OR_IVW_ = 1.29, 95%CI = 0.85 – 1.95; Table 3). Single SNP analyses resulted in a similar effect estimate for the variant in the *AMH* locus (OR_IVW_ = 1.27, 95%CI = 0.64 – 2.56), risk increasing effects for the SNPs in the *TEX41* and *MCM8* loci, and a risk decreasing effect of the variant in the *CDCA7* locus (Table 3). The heterogeneity test did not suggest heterogeneous effects of the individual SNPs (p = 0.30), most likely because of the high uncertainty in individual SNP estimates, but again interpretation of this p-value is limited with only four SNPs. Leave-one-out sensitivity analyses indicated rs11683493 (*CDCA7* locus) affected the IVW estimate most, and that exclusion of this variant resulted in a positive association (OR_IVW_ = 1.53, 95%CI = 1.01 – 2.33) (Figure S13).

## Discussion

We identified four loci for circulating AMH levels in women of European ancestry. In addition to confirming a previously reported signal in the *MCM8* locus, we discovered three new signals in and near the *AMH, TEX41* and *CDCA7* genes. In total, 35 genes were prioritized for these loci based on physical position, eQTL mapping and chromatin interactions, but pathway analyses did not reveal enrichments of gene-sets, tissues or cell types for genes annotated to suggestive associated SNPs. Genetic correlation analyses supported a shared genetic architecture between AMH levels and age at menopause. Exploratory MR analyses did not provide strong evidence of a causal effect of circulating AMH on breast cancer and PCOS.

We confirmed the association between rs16991615 and circulating AMH levels, previously reported by Ruth et al.^8^. This SNP is a missense variant located in exon 9 of the *MCM8* gene, rendering *MCM8* the most likely causal gene at this locus. In humans, *MCM8* plays a role in in homologous recombination, which is critical for DNA repair.^42^ Previous studies have linked *MCM8* deficiency to premature ovarian failure and infertility, but also to cancer development.^39; 43; 44^ Associations between rs16991615 and age at menopause^45^ and number of ovarian follicles^46^ have also been reported, which suggests that this locus is associated with circulating AMH levels because of its influence on the number of antral follicles.

Our GWAS study is the first AMH GWAS that identified a missense SNP (rs10417628) in the *AMH* gene in women. A previous AMH GWAS including adolescents from ALSPAC identified three SNPs in the *AMH* gene that were only significantly associated with AMH levels in male adolescents, and of which one (rs2385821) was in moderate LD with our lead SNP rs10417628 (R^2^ = 0.55).^6^ However, approximate conditional and joint analyses suggested that these variants represent the same signal at the *AMH* locus. Although identification of a genetic variant in the gene encoding for AMH itself suggests that we reveal an actual signal for circulating AMH concentrations, a recent case report suggests that the amino acid substitution corresponding to rs10417628 reduces AMH detection by the picoAMH assay from Ansh Labs without influencing AMH bioactivity.^47^ We sought to verify this finding in a subsample of the Doetinchem Cohort Study, for which AMH was measured using both the picoAMH and the less sensitive Gen II assay from Beckman Coulter. For the only woman who was estimated to be homozygous for the T allele (dosage T allele = 1.9, age at measurement = 28.3 years), AMH levels were indeed undetectable using the picoAMH assay, whereas circulating AMH levels were detected using the Gen II assay (318 pg/mL). In addition, median AMH levels measured using the Gen II assay were less different between women homozygous for the reference allele and heterozygous women (median AMH levels_homozygousrefallele_ = 953.0 pg/mL, IQR: 428.0 - 1999.0; median AMH levels_heterozygous_ = 848.0 pg/mL, IQR: 509.0 - 1310.0), compared to AMH levels measured using the picoAMH assay (median AMH levels_homozygousrefallele_ = 1485.9 pg/mL, IQR: 704.4 - 3150.0; median AMH levels_heterozygous_ = 811.0 pg/mL, IQR: 462.3 - 1480.9). For ALSPAC, which also used the Gen II assay to measure AMH, the distribution of AMH levels was similar across adult women homozygous for the reference allele and heterozygous women as well. However, in the ALSPAC daughters median AMH levels were clearly higher in adolescents homozygous for the reference allele compared to heterozygous adolescents. Among the ALSPAC participants, only one adolescent was homozygous for the T allele, but her AMH levels could not be shared due to disclosure risk. Because of the lack of publicly available information on the antibodies and conformational epitopes of the Gen II assay, and the limited and inconsistent evidence in the current study, we do not want to draw any definite conclusions about this yet.

For the associated loci on chromosome 2 it is more challenging to assign possible causal genes, as *TEX41* is a long noncoding RNA and the SNP in the *CDCA7* locus was located in an intergenic region. Gene mapping based on chromatin interactions with *TEX41* highlighted several genes, including the long non-coding RNA *ZEB2-AS1* (ZEB2 antisense RNA 1). *ZEB2-AS1* up-regulates expression of the protein ZEB2.^48^ ZEB2 (also known as SIP1) inhibits signal transduction in TGF-β and BMP signalling through interaction with ligand-activated SMAD proteins.^49; 50^ Among other BMP proteins, BMP4 has been reported to regulate AMH expression through activation of SMAD proteins.^51; 52^ Based on identification of *BMP4* in our gene-based association analysis and its prioritization by DEPICT, we hypothesize that BMP4 induced AMH expression may be regulated by ZEB2 interaction. However, fundamental laboratory research is needed to prove this.

*CDCA7* (also known as *JPO1*) is a direct target gene of the transcription factor *MYC* and is involved in apoptosis.^53^ Functional annotation did not map any genes to this locus and thus the mechanism through which this locus affects circulating AMH levels remains to be elucidated. Also for *EIF4EBP1*, and *GAB2*, which were significantly associated with AMH levels in the gene-based association analysis, it is not yet clear if and how these genes influence AMH expression. A target gene of *EIF4EBP1* has been previously highlighted as candidate gene for age at menopause^54^, whereas *GAB2* is involved in FSH signalling^55^ and has been identified as a potential causal gene for age at menarche.^56^ Altogether these results suggest that these genes play a role in reproductive functioning and reproductive aging, potentially by affecting the number of antral follicles, which are the main producers of AMH in women.^2^ Ideally, future studies should explore whether the observed genetic associations may merely reflect the size of the ovarian reserve, through adjusting analyses for antral follicle count. Such analyses would also show if we can actually use the identified variants are instruments for circulating AMH levels itself or for the quantity of antral follicles in MR analyses.

We did not find support for a causal effect of circulating AMH levels on breast cancer and PCOS risk in our exploratory MR analyses. To be valid genetic instruments for MR, SNPs have to fulfil the following three criteria^57^: (1) SNPs have to be associated with the circulating AMH levels; (2) SNPs cannot be associated with confounders of the studied AMH-outcome associations, and (3) SNPs cannot influence the outcomes through mechanisms that do not involve circulating AMH levels. Because rs10417628 in the *AMH* gene potentially reflects AMH detection instead of AMH expression, analyses including this variant should be interpreted with caution. However, leave-one-out analyses excluding this variant did not affect IVW MR estimates. Based on the function of genes mapped to the loci on chromosomes 20 and 2, it is likely that these variants affect breast cancer and PCOS risk through mechanisms independent of AMH (e.g. DNA replication and apoptosis), in particular the *MCM8* locus, which has also been identified in breast cancer GWAS.^39^ Due to the limited number of identified lead SNPs it was not possible to assess if our results were indeed biased by horizontal pleiotropy. Furthermore, weak instrument bias may still have biased MR results towards the null, since the F statistics may be overestimated in this GWAS (853.9 for breast cancer, and 422.4 for PCOS). Consequently, we should be cautious about excluding a causal effect of AMH on the studied outcomes.

Previous research suggests that AMH levels in females rise during puberty, until the mid to late twenties, and after that decrease until menopause.^11; 58^ Based on these observations and the differences in both age and AMH distributions between the ALSPAC adolescents and other participants, we performed a sensitivity analysis excluding the adolescents from ALSPAC. This analysis revealed an additional locus on chromosome 5 (rs116090962, nearest gene: *CTB-99A3*.*1*), although we could not find clues for its association with circulating AMH levels in adult women only, nor with AMH levels in general. Study-specific betas revealed an opposite effect for the *MCM8* locus and a minimal effect for the *CDCA7* locus in adolescents compared to effects in adult women. A larger GWAS including older adolescents and a larger proportion of females aged 20 to 40, would be required to reveal potential gene-age interactions explain variation in AMH expression.

The main strengths of the current GWAS meta-analysis are its size, which is twice the size of the previous GWAS meta-analysis, and its larger proportion of women of early-reproductive age. Given AMH’s function in ovarian follicle development, circulating levels and variation in AMH levels decrease with age. As a result, statistical power to identify genetic variants for circulating AMH increases if younger women are included. Still, our sample size remains relatively small for a GWAS, and future larger studies may lead to the detection of additional variants for circulating AMH levels. This is supported by our chip heritability estimate of 15% (se = 7%), which indicates that there are likely more SNPs that contribute to variability in AMH levels. Identification of additional genetic variants will also facilitate increased power to identify pathways and tissues enriched for genes involved in AMH expression. A second limitation of this study is potential overlap in participants between the current AMH GWAS and the GWAS for breast cancer^39^ (maximum n = 1,459; 20.7% of current study) and PCOS^40^ (maximum n = 225; 3.2% of current study). Overlap in participants in two-sample MR analyses may bias effect estimates and inflate Type 1 error rates.^59^

In conclusion, we replicated the previously reported association with the *MCM8* locus and identified 3 novel loci for circulating AMH levels in women, including the *AMH* locus. The strongest signal in this locus possibly affects AMH detection by specific assays rather than AMH bioactivity, but further research is required to confirm this hypothesis. Genes mapped to the *MCM8, TEX41* and *CDCA7* loci are involved in the cell cycle and processes like DNA replication and apoptosis. The mechanism underlying their associations with AMH may affect the size of the ovarian follicle pool. MR analyses did not support a causal effect of AMH on breast cancer and PCOS, but these finding should be interpreted with caution because we could not robustly explore how valid the instruments were and weak instrument bias may have biased estimates towards the null.

## Data Availability

Code generated for this study is available at Github. The full AMH GWAS summary statistics will made available through the GWAS catalog (https://www.ebi.ac.uk/gwas/).

https://github.com/reneemgverdiesen/AMH_GWAS

## Supplemental Data

Document S1: Supplemental Methods.

Document S2: Figures S1 – S13.

Document S3: Tables S1 – S11.

## Acknowledgments

The Generations Study thank Breast Cancer Now and the Institute of Cancer Research for support and funding of the Generations Study, and the Study participants, Study staff, and the doctors, nurses and other health care staff and data providers who have contributed to the Study. ICR acknowledges NHS funding to the NIHR Biomedical Research Centre. We would like to thank the participants and staff of the Nurses’ Health Study and the Nurses’ Health Study II.

For the Doetinchem Cohort Study we thank dr HSJ Picavet, A Blokstra MSc and Petra Vissink for their crucial role in data collection and data management.

For the ALSPAC Cohort, we are extremely grateful to all the families who took part in this study, the midwives for their help in recruiting them, and the whole ALSPAC team, which includes interviewers, computer and laboratory technicians, clerical workers, research scientists, volunteers, managers, receptionists and nurses.

SWAN thanks the study staff at each site and all the women who participated in SWAN. <u>Clinical Centers:</u> University of Michigan, Ann Arbor – Siobán Harlow, PI 2011 – present, MaryFran Sowers, PI 1994-2011; Massachusetts General Hospital, Boston, MA – Joel Finkelstein, PI 1999 – present; Robert Neer, PI 1994 – 1999; Rush University, Rush University Medical Center, Chicago, IL – Howard Kravitz, PI 2009 – present; Lynda Powell, PI 1994 – 2009; University of California, Davis/Kaiser – Ellen Gold, PI; University of California, Los Angeles – Gail Greendale, PI; Albert Einstein College of Medicine, Bronx, NY – Carol Derby, PI 2011 – present, Rachel Wildman, PI 2010 – 2011; Nanette Santoro, PI 2004 – 2010; University of Medicine and Dentistry – New Jersey Medical School, Newark – Gerson Weiss, PI 1994 – 2004; and the University of Pittsburgh, Pittsburgh, PA – Karen Matthews, PI. <u>NIH Program Office:</u> National Institute on Aging, Bethesda, MD – Chhanda Dutta 2016-present; Winifred Rossi 2012–2016; Sherry Sherman 1994 – 2012; Marcia Ory 1994 – 2001; National Institute of Nursing Research, Bethesda, MD – Program Officers. <u>Central Laboratory:</u> University of Michigan, Ann Arbor – Daniel McConnell (Central Ligand Assay Satellite Services). <u>SWAN Repository</u>: University of Michigan, Ann Arbor – Siobán Harlow 2013 - Present; Dan McConnell 2011 - 2013; MaryFran Sowers 2000 – 2011. <u>Coordinating Center:</u> University of Pittsburgh, Pittsburgh, PA – Maria Mori Brooks, PI 2012 - present; Kim Sutton-Tyrrell, PI 2001 – 2012; New England Research Institutes, Watertown, MA - Sonja McKinlay, PI 1995 – 2001. <u>Steering Committee</u>: Susan Johnson, Current Chair, Chris Gallagher, Former Chair.

The Sister Study acknowledges the contributions of Jack A. Taylor, M.D., Ph.D., Molecular & Genetic Epidemiology Group, Epidemiology Branch, National Institute of Environmental Health Sciences, Durham, NC who secured initial funding for the Sister Study GWAS, and Hazel B. Nichols. Ph.D., Associate Professor, Department of Epidemiology, Gillings School of Global Public Health who was the PI of the grant that allowed the AMH assays for the case-control study of AMH and breast cancer in premenopausal women.

## Web Resources

ALSPAC: http://www.bristol.ac.uk/alspac/

SWAN: https://www.swanstudy.org/

EASYQC: https://www.uni-regensburg.de/medizin/epidemiologie-praeventivmedizin/genetische-epidemiologie/software/

METAL: http://csg.sph.umich.edu/abecasis/metal/

GCTA: https://cnsgenomics.com/software/gcta/

FUMA: https://fuma.ctglab.nl/

DEPICT: https://github.com/perslab/depict

LD Hub: http://ldsc.broadinstitute.org/

## Data and Code Availability

Code generated for this study is available at Github [https://github.com/reneemgverdiesen/AMH_GWAS]. The full AMH GWAS summary statistics will made available through the GWAS catalog (https://www.ebi.ac.uk/gwas/)

