## Supplemental Figures for "Genome-wide association study meta-analysis identifies three novel loci for circulating anti-Müllerian hormone levels in women"


| | |
|---|---|
| **Figure S1** | Meta-level QC - SE-N plot |
| **Figure S2** | Meta-level QC - P-Z plots |
| **Figure S3** | Meta-level QC - Allele frequency plots |
| **Figure S4** | Meta-level QC - QQ plots |
| **Figure S5** | Meta-level QC - Lambda-N plot |
| **Figure S6** | QQ plot for meta-analysis ALSPAC mothers and daughters only |
| **Figure S7** | QQ plot for meta-analysis all studies |
| **Figure S8** | Regional association plots for genome-wide significant loci |
| **Figure S9** | Manhattan plot of gene-based genome-wide association results for inverse normally transformed AMH in women |
| **Figure S10** | Scatter plot of genetic associations with breast cancer against genetic associations with circulating AMH |
| **Figure S11** | Estimates leave-one-out analyses for the association between circulating AMH and risk of breast cancer |
| **Figure S12** | Scatter plot of genetic associations with PCOS against genetic associations with circulating AMH |
| **Figure S13** | Estimates leave-one-out analyses for the association between circulating AMH and risk of PCOS |

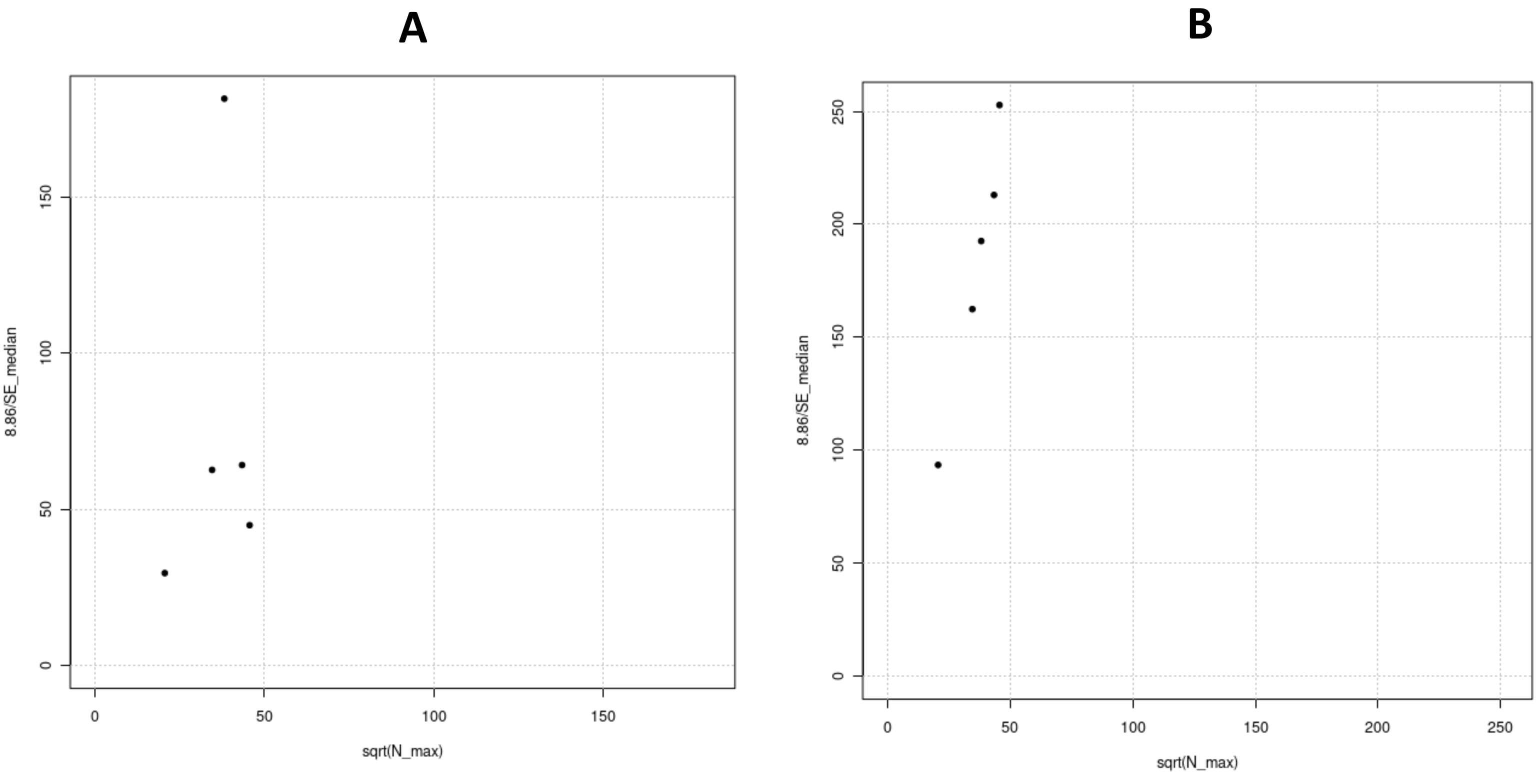


**Figure S1: SE-N plot to detect issues with trait transformations.**

The individual points represent, for each included study, the inverse of the median standard error of the beta estimates across all SNPs against the square root of the sample size. If there are no issues with trait transformations the data points are more or less on a straight line. (A) Before filtering on minor allele frequency; the data point for the GWAS by Ruth et al. appears an outlier compared to the data points of the other studies. (B) After filtering on minor allele frequency previous (MAF > 1%); data points are more or less on a straight line. Plots were created using the EasyQC R-package.[1]

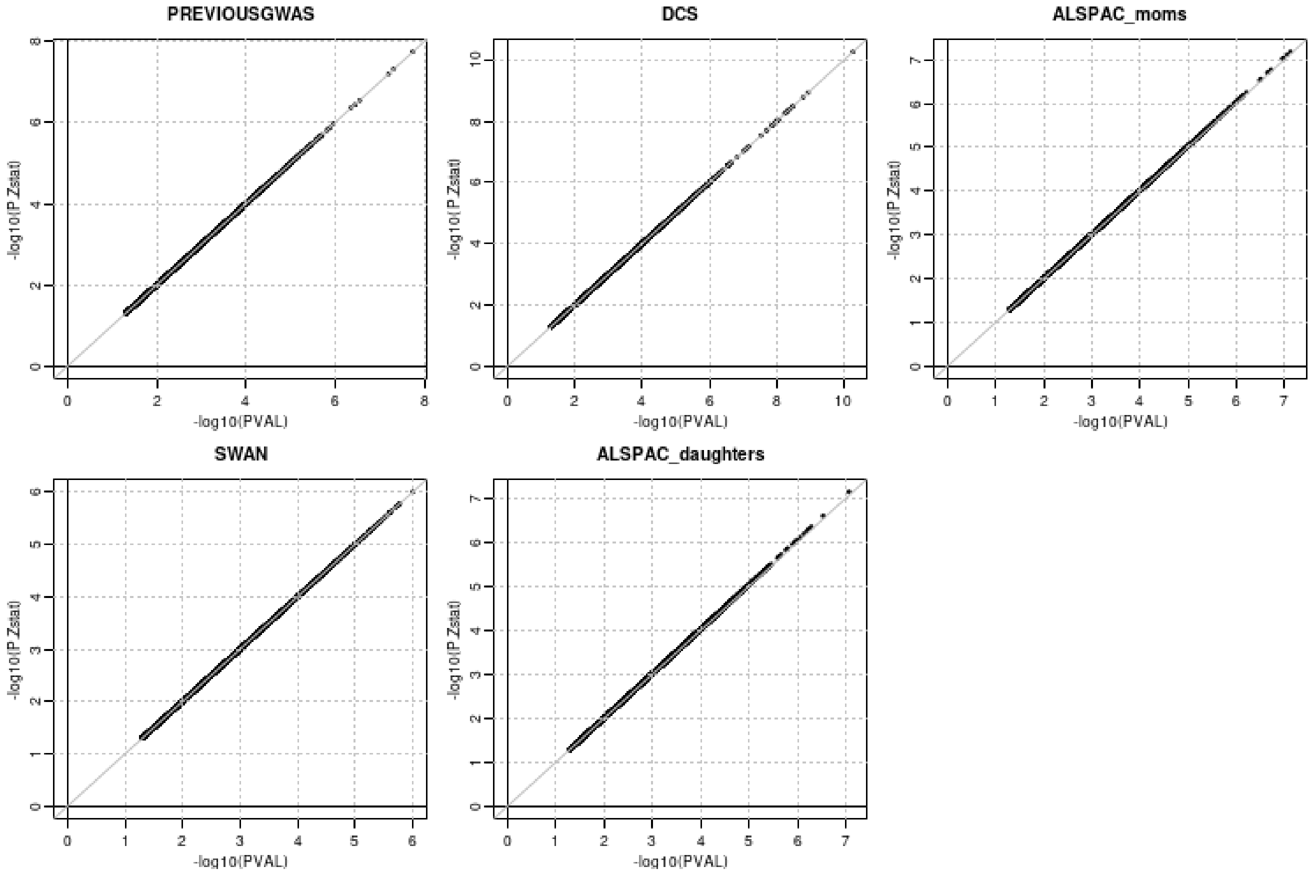


**Figure S2: P-Z plots to detect analytical issues with beta, standard errors and P-values.**

For each participating study, observed p-values are compared with P-values calculated from the Z-statistics based on the observed beta-estimates and standard errors. A straight line indicates that observed and calculated p-values are in agreement, and thus no analytical issues with betas standard errors and p-values are present for the included studies. Plots were created using the EasyQC R-package.[1]

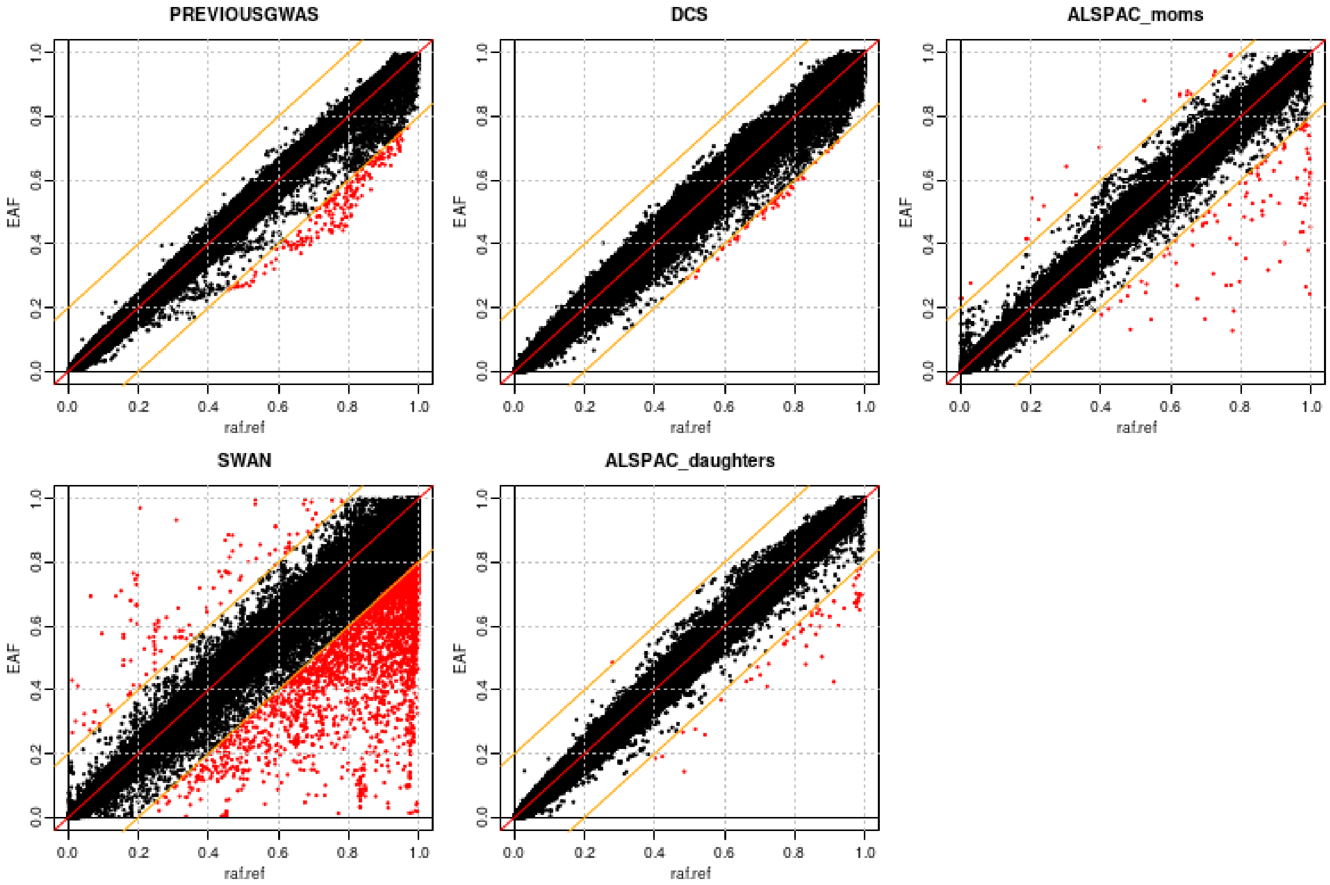


**Figure S3: Allele frequency plots to check for issues with allele frequency.**

For each included study, effect allele frequencies (EAF) are plotted against the HRC reference panel. For the GWAS by Ruth et al., the Doetinchem Cohort Study, and ALSPAC data are very consistent with the reference panel. Although the plot for SWAN indicates various differences with the HRC panel, none of the previously described specific patterns for systematic deviations are observed. Most likely the SWAN population have a somewhat different ancestry than the reference. Plots were created using the EasyQC R-package.[1]

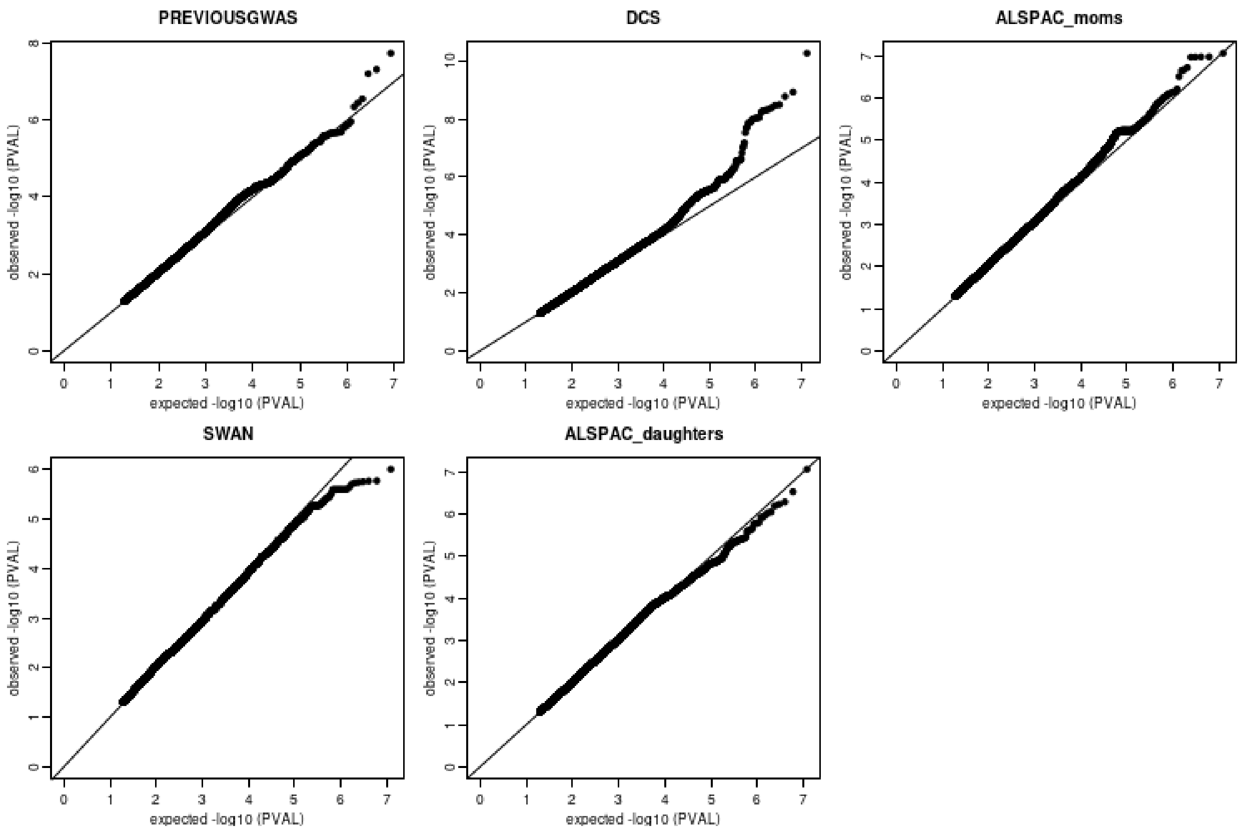


**Figure S4 QQ plots per included study before removal of SNPs with MAF < 1% and poor imputation quality.**

For each study, observed –log10 p-values for each SNP are plotted against expected –log10 p-values from a theoretical $\chi^2$ distribution. Plots were created using the EasyQC R-package.[1]

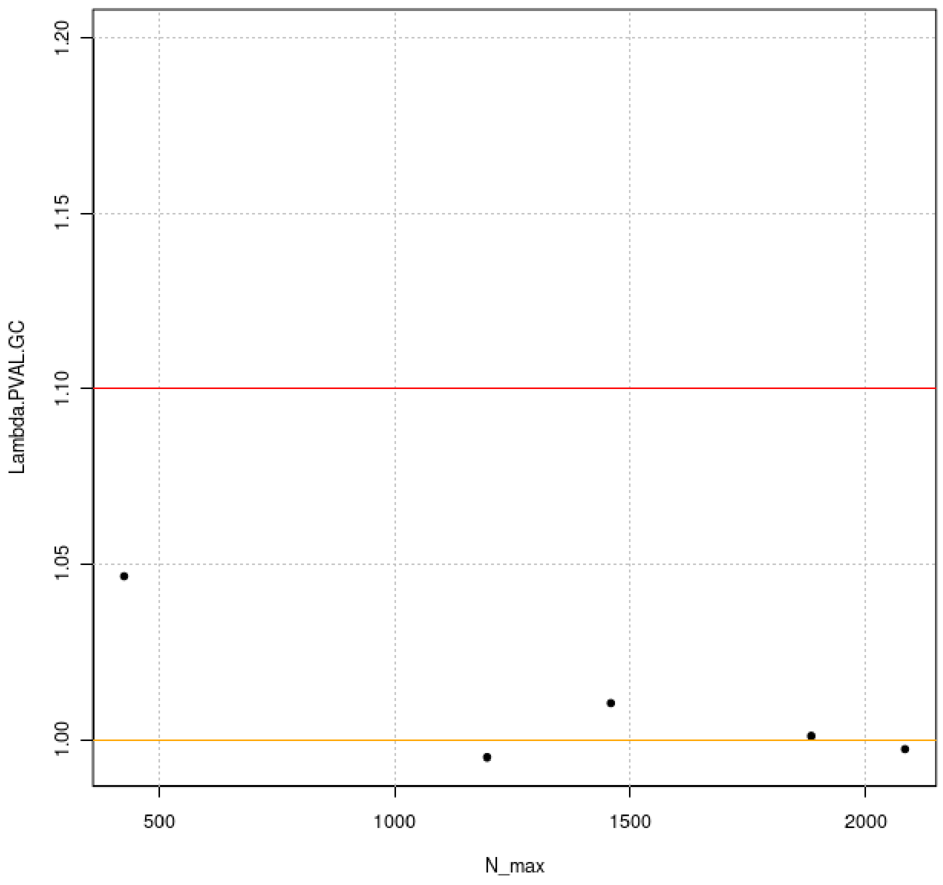


**Figure S5: Lambda-N plot to reveal issues with population stratification.**

For each study, the lambda for genomic control is plotted against the maximum sample size. The orange line indicates the optimal lambda; $\lambda_{GC} = 1.0$. The red line indicates the threshold for values that indicate problems with population stratification; $\lambda_{GC} = 1.1$. This plot suggests that none of the included studies has population stratification issues. Plot was created using the EasyQC R-package.[1]

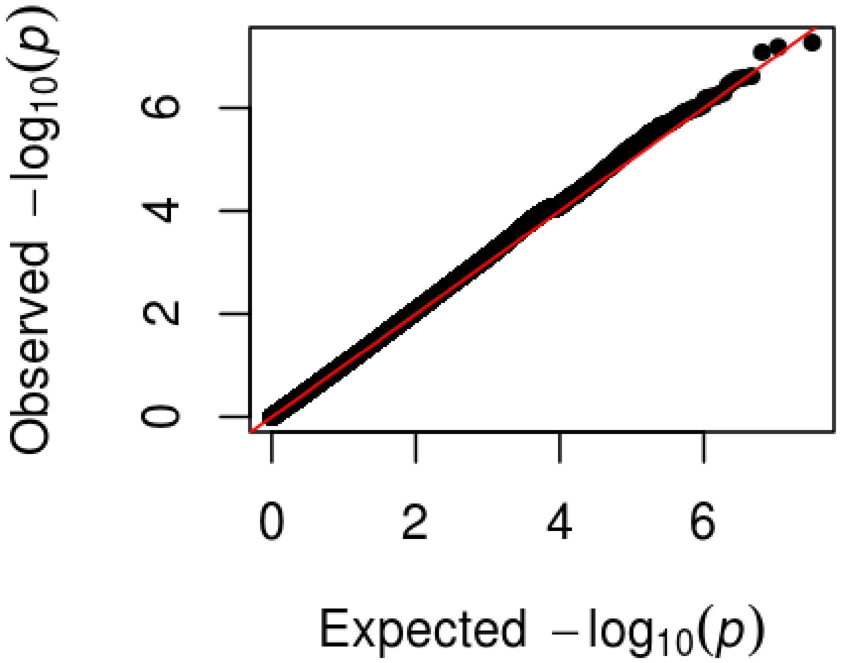


**Figure S6: QQ plot of meta-analysis p-values of ALSPAC mothers and daughters only.**

Observed –log10 p-values from the meta-analysis including only ALSPAC mothers and daughters are plotted against expected –log10 p-values from a theoretical $\chi^2$ distribution. Corresponding lambda is 1.009.

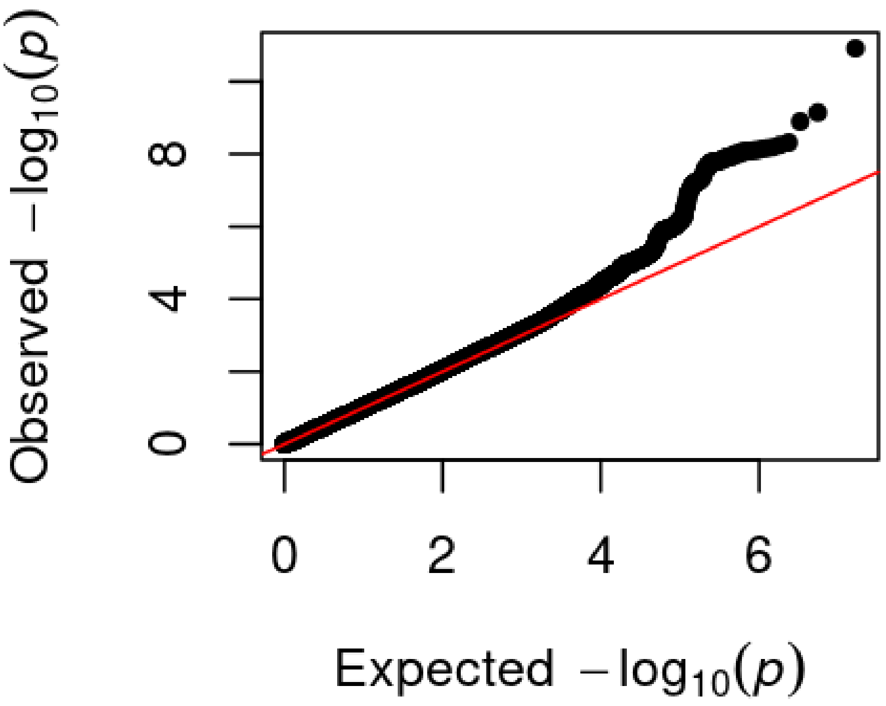


**Figure S7: QQ plot of meta-analysis p-values for inverse normally transformed AMH in women.**

Observed –log10 p-values for each of the 8,298,138 SNPs included in the meta-analysis are plotted against expected –log10 p-values from a theoretical $\chi^2$ distribution. Corresponding lambda is 1.006.

**A**

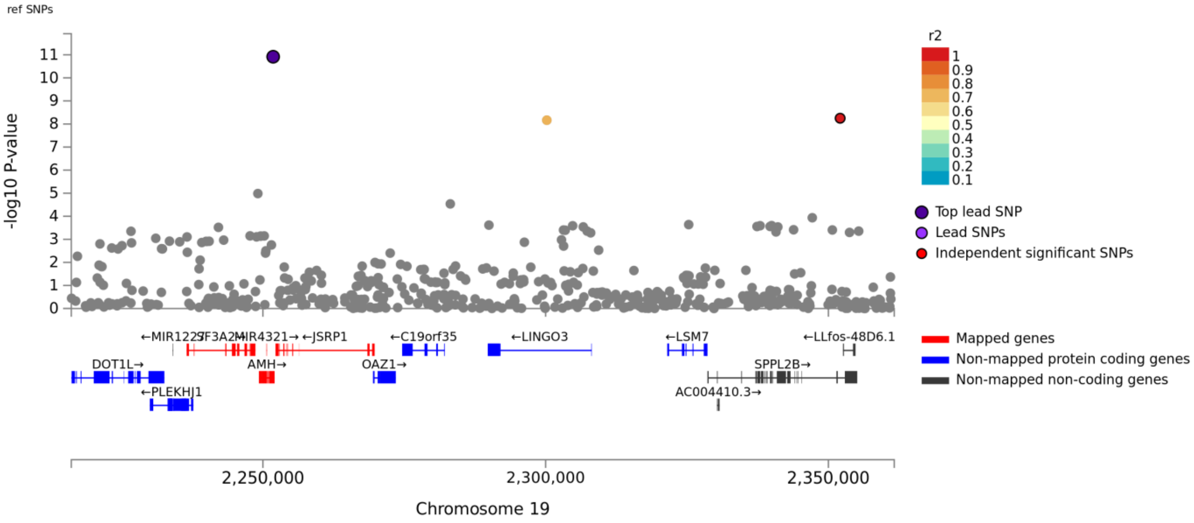
ref SNPs
-log10 P-value
r2
Top lead SNP
Lead SNPs
Independent significant SNPs
←MIR1227
SF3A2
←MIR4321→
←JSRP1
←C19orf35
←LINGO3
←LSM7
←LLfos-48D6.1
DOT1L→
AMH→
OAZ1→
SPPL2B→
←PLEKHJ1
AC004410.3→
Mapped genes
Non-mapped protein coding genes
Non-mapped non-coding genes
2,250,000
2,300,000
2,350,000
Chromosome 19

**B**

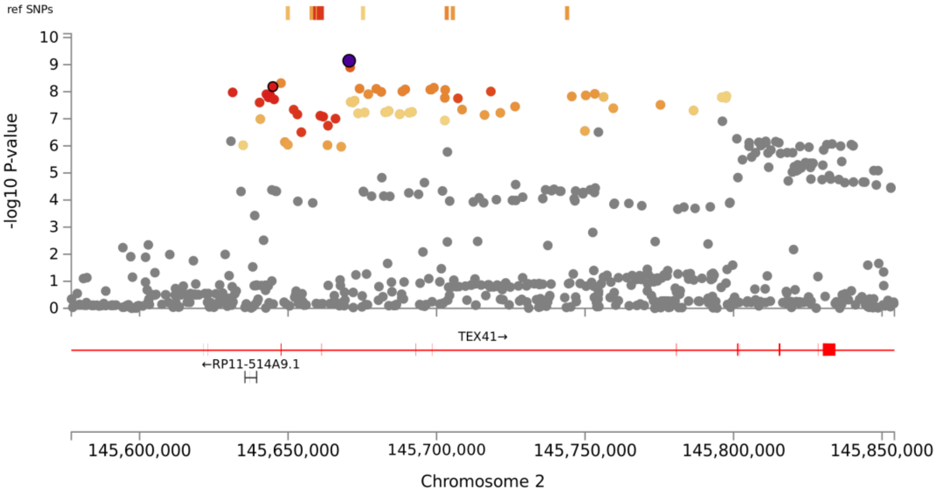
ref SNPs
-log10 P-value
TEX41→
←RP11-514A9.1
145,600,000
145,650,000
145,700,000
145,750,000
145,800,000
145,850,000
Chromosome 2

**C**

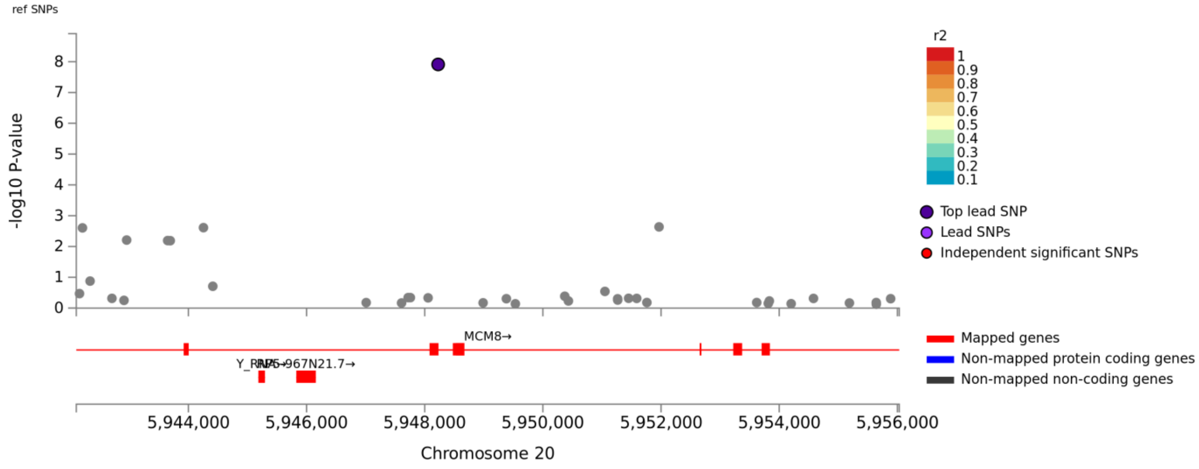


**D**

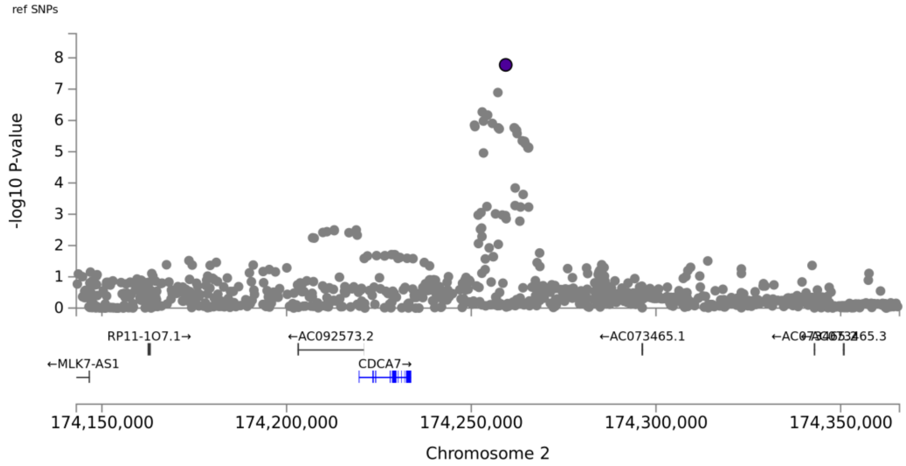


**Figure S8: Regional association plots for genome-wide significant loci for inverse normally transformed AMH in women.**

Regional plots for the *AMH* locus (panel A), *TEX41* locus (panel B), *MCM8* locus (panel C), and *CDCA7* locus (panel D) show SNPs plotted by their position and –log10 P-value for association with inverse normally transformed AMH. Nearby genes are depicted below each plot. Plots were created using FUMA.[2]

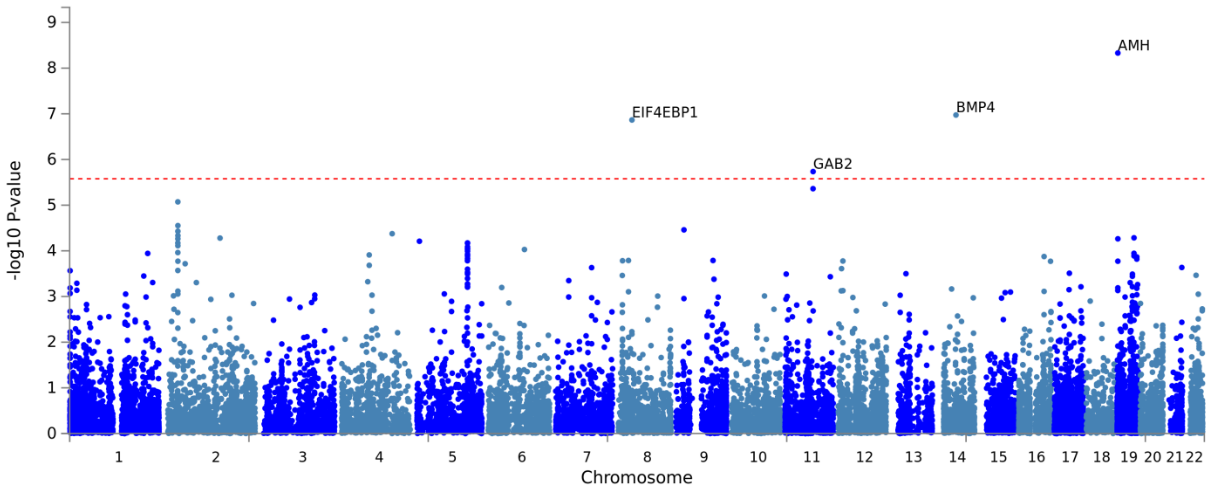


**Figure S9: Manhattan plot of gene-based genome-wide association results for inverse normally transformed AMH in women.**

Plot was created using FUMA.[2]

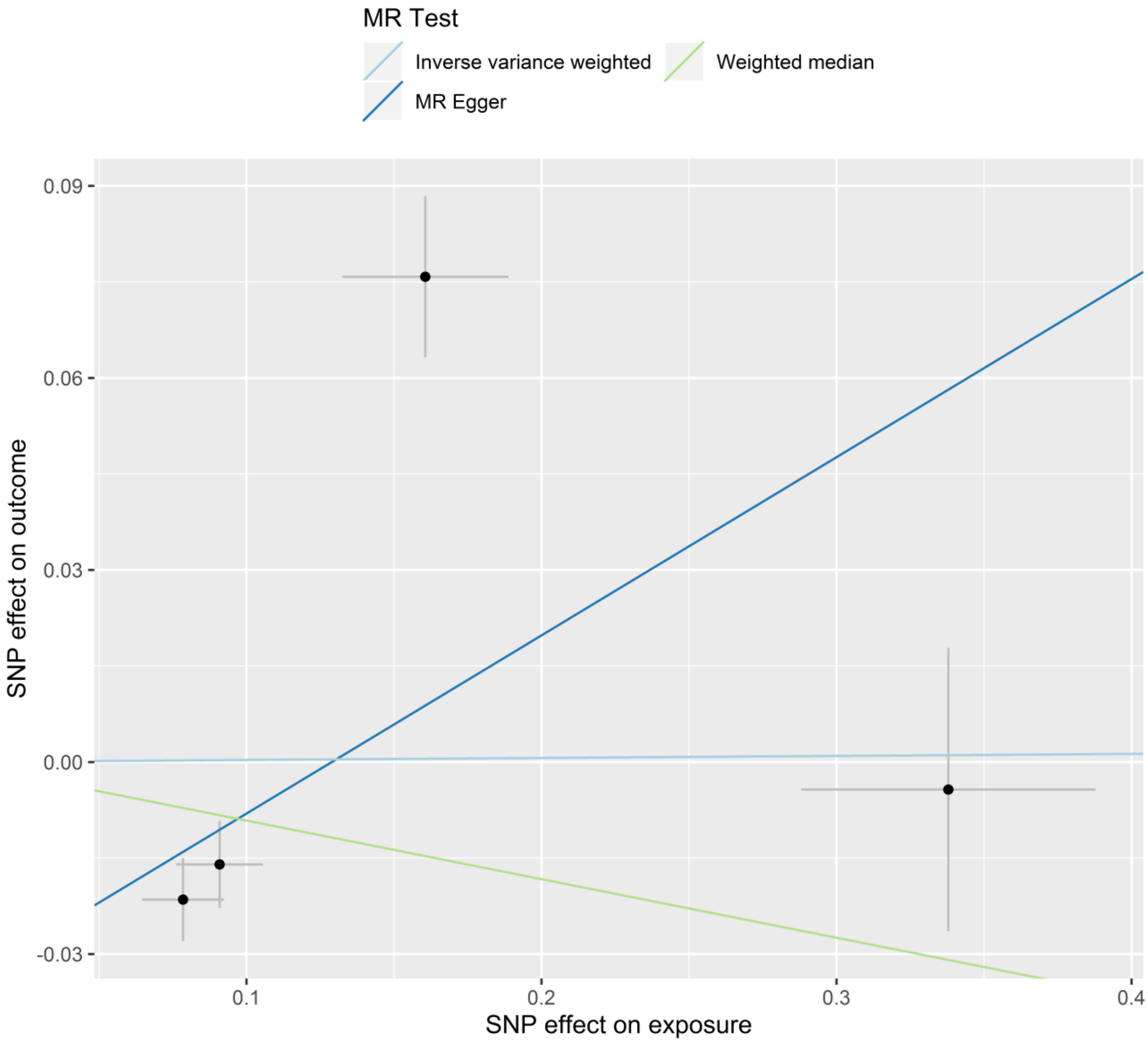


**Figure S10: Scatter plot of genetic associations with breast cancer (vertical axis) against genetic associations with circulating AMH (horizontal axis).**

Plot was created using the TwoSampleMR R-package.[3]

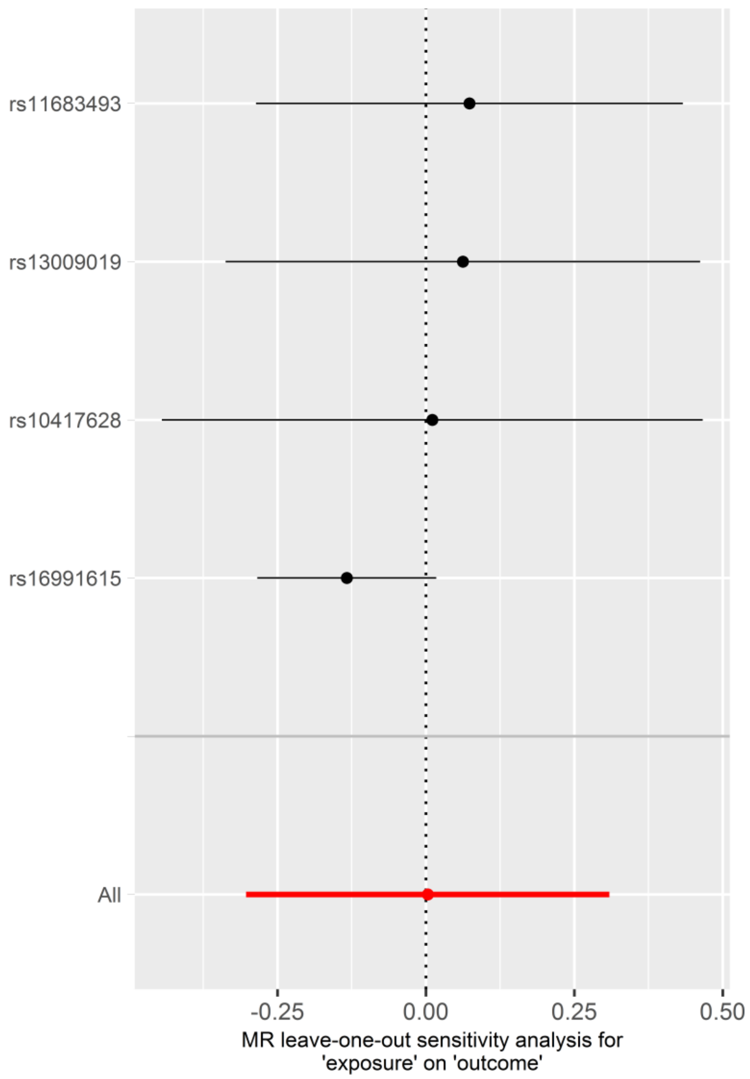


**Figure S11: Estimates leave-one-out analyses for the association between circulating AMH and risk of breast cancer.**

Plot was created using the TwoSampleMR R-package.[3]

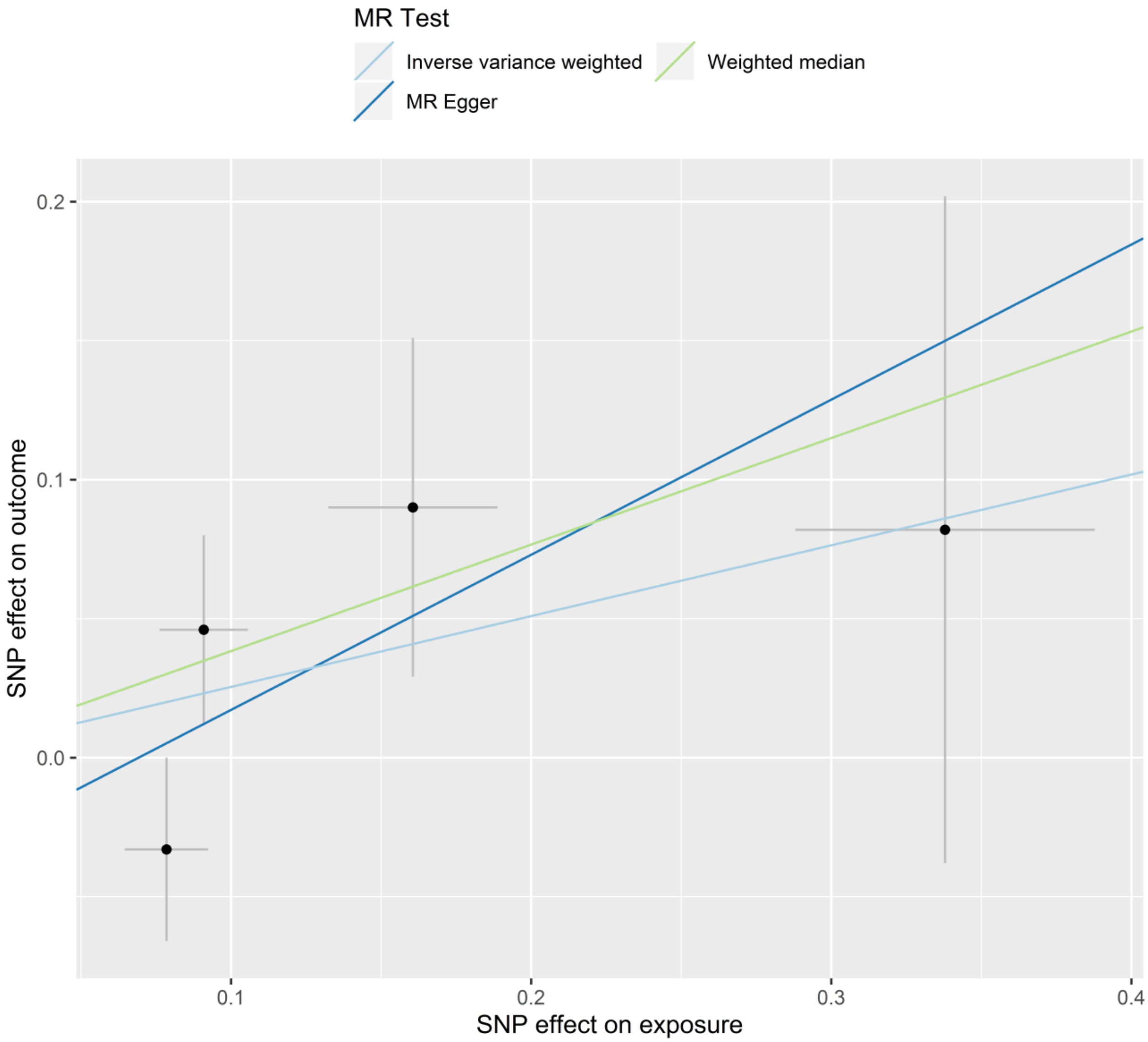


**Figure S12: Scatter plot of genetic associations with PCOS (vertical axis) against genetic associations with circulating AMH (horizontal axis).**

Plot was created using the TwoSampleMR R-package.[3]

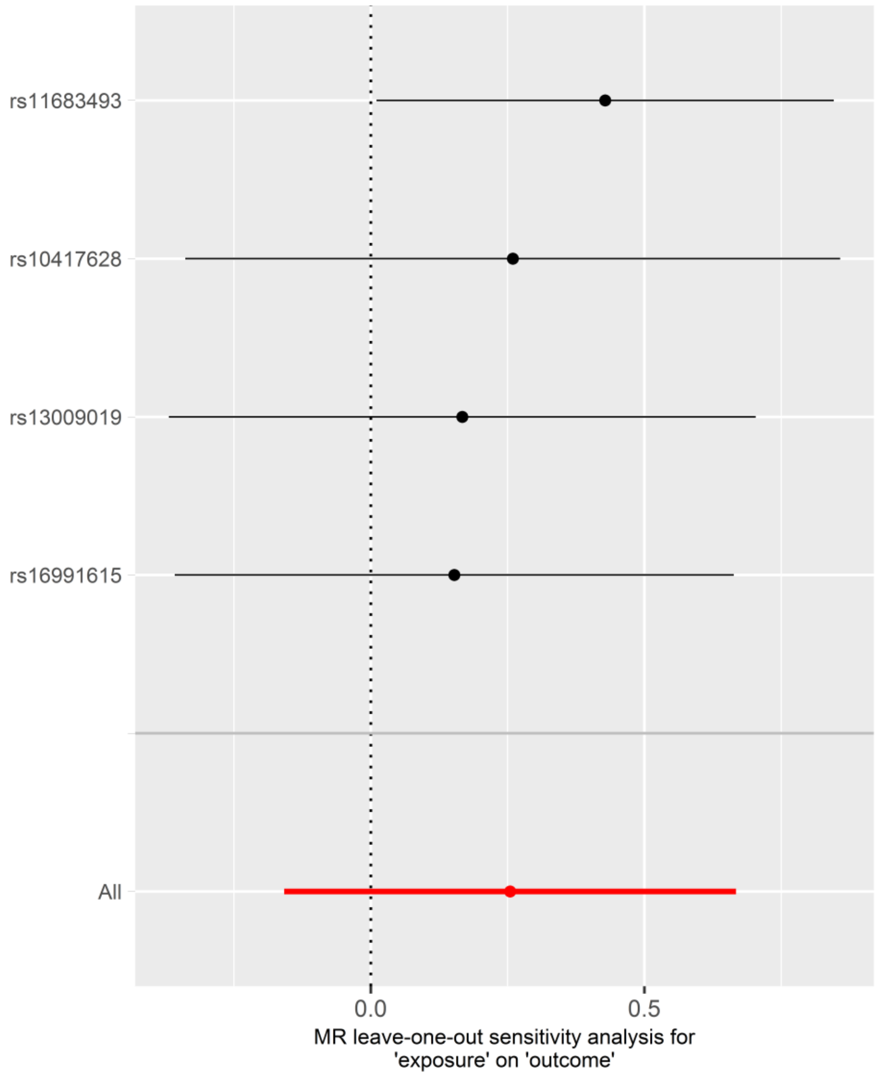


**Figure S13: Estimates leave-one-out analyses for the association between circulating AMH and risk of PCOS.**

Plot was created using the TwoSampleMR R-package.[3]
