## Supplemental Methods for "Genome-wide association study meta-analysis identifies three novel loci for circulating anti-Müllerian hormone levels in women"

#### Study population

*Studies included in the previous GWAS study: Generations Study, Sister Study, Nurses' Health Study and Nurses' Health Study II*

We included AMH GWAS summary statistics from the most recent previous AMH GWAS analysis.[1] We requested summary statistics excluding data from ALSPAC, resulting in meta-analysis summary statistics for the Generations Study (n = 379, median age: 44 years, IQR: 40, 48), Sister Study (n = 438, median age: 48 years, IQR: 45, 51), Nurses' Health Studies (n = 642, median age: 44, IQR: 41, 47). These population-based cohort studies have been described in more detail previously.[2-5] Details on the premenopausal women included in the previous AMH GWAS by Ruth et al. are presented in Supplementary Table 1 and are described into more detail elsewhere.[1; 6]

*Doetinchem Cohort Study*

The Doetinchem Cohort Study is an ongoing prospective cohort study that included 3,641 men and 4,128 women, aged 20-59 years at recruitment, who were randomly selected from the municipal register of Doetinchem, The Netherlands, between 1987 and 1991. Every five years, study participants are invited for a follow-up visit, during which physical examinations and extensive questionnaires are completed, and blood samples are collected. The Doetinchem Cohort Study received ethical approval from the Medical Ethics Committee of The Netherlands Institution of Applied Scientific Research and all study participants signed an informed consent prior to study inclusion. For more details see previous reports.[7; 8] Details on the included participants are presented in Supplementary Table 1.

*Avon Longitudinal Study of Parents and Children*

The Avon Longitudinal Study of Parents and Children (ALSPAC) is a longitudinal birth cohort, which has been described in detail elsewhere.[9; 10] In short, 14,541 women who were expected to give birth between 1st April 1991 and 31st December 1992 from the South West of England were enrolled in ALSPAC between 1990 and 1992.[10] Initially, 14,676 fetuses were included in ALSPAC. When the oldest children were approximately 7 years old, an attempt was made to bolster the initial sample with eligible cases who had failed to join the study originally. The total sample size for analyses using any data collected after the age of seven is therefore 15,454 pregnancies, resulting in 15,589 foetuses. Of these 14,901 were alive at 1 year of age.

Ethical approval for the study was obtained from the Avon Longitudinal Study of Parents and Children Ethics and Law Committee and the Local Research Ethics Committees. Written informed consent was obtained from all adult participants in the study. Consent for biological samples has been collected in accordance with the Human Tissue Act (2004). Please note that the study website contains details of all the data that is available through a fully searchable data dictionary and variable search tool and reference the following webpage: <http://www.bristol.ac.uk/alspac/researchers/our-data/>. Details on the included participants are presented in Supplementary Table 1.

*Study of Women's Health Across the Nation*

The Study of Women's Health Across the Nation (SWAN) is a multi-site, multiracial/ethnic longitudinal study of women's health designed to describe the biological, behavioral, and psychosocial characteristics that occur during midlife and the menopausal transition. Briefly, the SWAN cohort was enrolled in 1996-97 and consists of 3302 community-based women from seven sites with data from five race/ethnic groups: Black (n=935), Chinese (n=250), Hispanic (n=286), Japanese (n=281), and White (n=1550). To be eligible for enrollment women had to be aged 42 to 52 years old, have an intact uterus and at least one ovary, have had a menstrual period in the previous three months, and not be taking hormones. Subsequently, 1,757 participants consented to provide genetic materials. Immortalized cell lines were developed successfully for 1,588, with 1,536 processed into distributable diluted, extracted DNA and 1,464 successfully genotyped. A total of 738 were of European ancestry, 425 of whom had AMH measures and contributed to this analysis. The study protocol was approved by the Institutional Review Boards at each study site. All participants provided written, informed consent at each visit. Details of SWAN are described elsewhere.[11] Details on the included participants are presented in Supplementary Table 1.

### Study-specific association analyses

The Doetinchem Cohort Study, ALSPAC, and SWAN performed association analyses based on a standardized analysis plan, which was distributed in advance. The analyses described in this analyses plan were in agreement with the analyses conducted for the previous AMH GWAS study.[1] All studies assumed an additive model and adjusted analyses for age at blood collection (years) and population stratification, either by including principal components (ALSPAC, SWAN) or a kinship matrix (Doetinchem Cohort Study, Generations Study, Sister Study, Nurses' Health Studies).

*Doetinchem Cohort Study*

We used rvtests[12] (version 20170210) to perform association analyses in the Doetinchem Cohort Study. Linear mixed model analyses were adjusted for age at blood collection and a kinship matrix was included to adjusted for cryptic relatedness. This kinship matrix was calculated using the vcf2kinship script provided by rvtests.

*Avon Longitudinal Study of Parents and Children*

Because of the large differences in both age and AMH distribution between the ALSPAC mothers and daughters, we considered it inappropriate to analyze them together using a linear mixed model method to correct for relatedness. Consequently, separate association analyses were conducted for the ALPSAC mothers and daughters. For both groups, linear regression analyses were performed in SNPTEST v2.5[13], and models were adjusted for age at blood collection and 10 principal components.

*Study of Women's Health Across the Nation*

Rvtests (version 20190205) was used to perform association testing in SWAN. Linear regression analyses were adjusted for age at blood draw and 10 principal components.

*Generations Study, Sister Study, Nurses' Health Study and Nurses' Health Study II*

For the Generations Study, Sister Study, and Nurses' Health Studies, linear mixed model association analyses were performed using GEMMA 0.94.1[14], which calculates a kinship matrix. Analyses were adjusted for age at blood draw. In the current meta-analysis we included summary statistics of the meta-analysis of these studies, which was performed using METAL[15], as described elsewhere.[1]

### File-level and meta-level QC prior to meta-analysis

Prior to meta-analysis, we performed file-level QC on all summary statistics files to clean and check the data, as described elsewhere.[16] File-level QC consisted of (1) removal of rows with missing data on alleles, p-values, betas, standard errors or allele frequency; (2) removal of rows with unrealistic values (e.g. p-values < 0 or > 1); (3) exclusion of monomorphic SNPs; (4) harmonizing alleles; and (5) removal of duplicated SNPs. Subsequently, we performed meta-level QC to identify potential study-specific problems following a previously published protocol.[16] Meta-level QC comprised creation of five plots: (1) SE-N plot, which reveals potential issues with trait transformation; (2) P-Z scatter plot, which reveals potential issues with betas, standard errors and p-values; (3) Allele frequency plot, to check for issues with allele frequencies or strand; (4) QQ plots, to assess genomic inflation, and (5) $\lambda_{GC}$ plot, also to assess genomic inflation. Both file-level and meta-level QC were performed using the R package EasyQC (v9.2).[16] No study-specific issues were identified through these QC procedures (Figure S1-S5).
